# Interstitial lung damage following COVID-19 hospitalisation: an interim analysis of the UKILD Post-COVID study

**DOI:** 10.1101/2022.03.10.22272081

**Authors:** I Stewart, J Jacob, PM George, PL Molyneaux, JC Porter, RJ Allen, JK Baillie, SL Barratt, P Beirne, SM Bianchi, JF Blaikley, J Chalmers, RC Chambers, N Chadhuri, C Coleman, G Collier, EK Denneny, A Docherty, O Elneima, RA Evans, L Fabbri, MA Gibbons, FV Gleeson, B Gooptu, NJ Greening, B Guillen Guio, IP Hall, NA Hanley, V Harris, EM Harrison, M Heightman, TE Hillman, A Horsley, L Houchen-Wolloff, I Jarrold, SR Johnson, MG Jones, F Khan, R Lawson, OC Leavy, N Lone, M Marks, H McAuley, P Mehta, E Omer, D Parekh, K Piper Hanley, M Platé, J Pearl, K Poinasamy, JK Quint, B Raman, M Richardson, P Rivera-Ortega, LC Saunders, R Saunders, MG Semple, M Sereno, A Shikotra, AJ Simpson, A Singapuri, DJF Smith, M Spears, LG Spencer, S Stanel, D Thickett, AAR Thompson, M Thorpe, R Thwaites, SLF Walsh, S Walker, ND Weatherley, M Weeks, JM Wild, DG Wootton, CE Brightling, LP Ho, LV Wain, RG Jenkins, the PHOSP-COVID Study Collaborative Group

**Affiliations:** National Heart & Lung Institute, Imperial College London; Respiratory Medicine, University College London; Royal Brompton and Harefield Clinical Group, Guy’s and St Thomas’ NHS Foundation Trust; University College London; University of Leicester; University of Edinburgh; North Bristol NHS Trust; Leeds Teaching Hospitals & University of Leeds; Sheffield Teaching Hospitals NHS Foundation Trust; University of Manchester; Manchester University NHS Foundation Trust; Ninewells Hospital and Medical School; University of Nottingham; University of Sheffield; University Hospitals of Leicester NHS Trust; Royal Devon and Exeter NHS Foundation Trust; Oxford University Hospitals NHS Foundation Trust; University of Oxford; University College London Hospital; Manchester University Hospitals NHS Foundation Trust; Asthma UK British Lung Foundation; Nottingham University Hospitals NHS Trust; Faculty of Medicine, University of Southampton; NIHR Southampton Biomedical Research Centre, University Hospital Southampton; Usher Institute, University of Edinburgh; London School of Hygeine and Tropical Medicine; University of Birmingham; University Hospitals Birmingham NHS Foundation Trust; British Lung Foundation; Liverpool University; Newcastle University; Perth Royal Infirmary, NHS Tayside; Liverpool University Hospitals NHS Foundation Trust; Sheffield Teaching NHS Foundation Trust; University of Liverpool

## Abstract

**Introduction:** Shared characteristics between COVID-19 and pulmonary fibrosis, including symptoms, genetic architecture, and circulating biomarkers, suggests interstitial lung disease (ILD) development may be associated with SARS-CoV-2 infection.

**Methods:** The UKILD Post-COVID study planned interim analysis was designed to stratify risk groups and estimate the prevalence of Post-COVID Interstitial Lung Damage (ILDam) using the Post-HOSPitalisation COVID-19 (PHOSP-COVID) Study. Demographics, radiological patterns and missing data were assessed descriptively. Bayes binomial regression was used to estimate the risk ratio of persistent lung damage >10% involvement in linked, clinically indicated CT scans. Indexing thresholds of percent predicted DLco, chest X-ray findings and severity of admission were used to generate risk strata. Number of cases within strata were used to estimate the amount of suspected Post-COVID ILDam.

**Results:** A total 3702 people were included in the UKILD interim cohort, 2406 completed an early follow-up research visit within 240 days of discharge and 1296 had follow-up through routine clinical review. We linked the cohort to 87 clinically indicated CTs with visually scored radiological patterns (median 119 days from discharge; interquartile range 83 to 155, max 240), of which 74 people had ILDam. ILDam was associated with abnormal chest X-ray (RR 1.21 95%CrI 1.05; 1.40), percent predicted DLco<80% (RR 1.25 95%CrI 1.00; 1.56) and severe admission (RR 1.27 95%CrI 1.07; 1.55). A risk index based on these features suggested 6.9% of the interim cohort had moderate to very-high risk of Post-COVID ILDam. Comparable radiological patterns were observed in repeat scans >90 days in a subset of participants.

**Conclusion:** These interim data highlight that ILDam was not uncommon in clinically indicated thoracic CT up to 8 months following SARS-CoV-2 hospitalisation. Whether the ILDam will progress to ILD is currently unknown, however health services should radiologically and physiologically monitor individuals who have Post-COVID ILDam risk factors.

## 1.0 Introduction

Long term symptoms of COVID-19 have been widely reported and can have a severe impact on quality of life, frequently characterized by chronic breathlessness.[1-3] Post-mortem studies on COVID-19 patients have highlighted diffuse parenchymal alterations, including alveolar damage, exudation, and development of pulmonary fibrosis, which may explain chronic respiratory symptoms in survivors.[4-6]

A number of studies have identified similarities between severe COVID-19 and idiopathic pulmonary fibrosis (IPF), an archetypal interstitial lung disease (ILD). These include shared genetic aetiology, [7, 8] circulating biomarkers, [9, 10] similarities in pulmonary function and radiological features.[11] This suggests that some survivors of COVID-19 may develop parenchymal abnormalities consistent with viral-associated ILD. To understand the potential risk of acute COVID-19 leading to interstitial lung damage (ILDam) or the development of longer term ILD and fibrosis, a longitudinal observational study of patients was planned, including individuals prospectively recruited following an admission to hospital with COVID-19 to the Post-HOSPitalisation COVID-19 (PHOSP-COVID) Study.[12].

To support clinical and research management, an interim analysis of the UKILD-Post COVID study was planned to estimate ILDam post hospitalisation after a minimum of one thousand participants had completed an early follow-up visit.[13] We present our approach for estimating the extent of suspected Post-COVID ILDam in hospital discharges across the UK, defined here as total lung involvement of reticulations and ground glass opacities >10% on clinically indicated CT.

## 2.0 Methods

### 2.1 Participants

This interim analysis was restricted to participants of the PHOSP-COVID study, a prospective longitudinal cohort study of adults discharged from National Health Service hospitals across the United Kingdom following admission for confirmed or clinical-diagnosed COVID-19. The PHOSP-COVID dataset includes a core set of demographics, tests of physical and pulmonary performance, symptom questionnaires, and biochemical tests, previously described in detail.[12] Participants were discharged by end of March 2021, interim data were collected up to October 2021.

Individuals withdrawing consent from PHOSP-COVID were excluded. Individuals being managed for an *a priori* diagnosed interstitial lung disease as recorded by site teams using hospital notes were identified by hand searches of comorbidities and excluded. This included, but was not limited to, recorded diagnoses of idiopathic pulmonary fibrosis, pulmonary sarcoidosis, asbestosis, hypersensitivity pneumonitis, cryptogenic organising pneumonia, combined emphysema and pulmonary fibrosis, and autoimmune related ILD.

### 2.2 Interim Study Design

For analyses, the UKILD interim cohort was restricted to PHOSP-COVID patients with clinically recorded data through routine follow-up (PHOSP-COVID Tier 1) and those with completed early research follow-up visits (PHOSP-COVID Tier 2), within 240 days of discharge. Clinically indicated thoracic CT scans were identified through the PHOSP-COVID study via linkage to a radiological database, all CT scans were requested at clinical discretion and may be unrelated to suspected ILDam or ILD. The presence of lung damage on volumetric CTs was described on a lobar basis. Across six lobes (the lingula was counted as a separate lobe) the percentage of reticulation and ground glass opacities were separately quantified by a single radiologist with over 10 years’ experience. Reticulation and ground glass opacities combined across six lobes was divided by 6, and the sum was used as the measure of total lung damage. The primary outcome was visually scored interstitial lung damage (ILDam) >10% lung involvement on CT.[14]

Index of Multiple Deprivation (IMD) was obtained using the patient’s postcode and presented as quintiles with areas of greatest deprivation in the first quintile. A modified WHO clinical progression scale was used to define the severity of admission (i. no supplemental oxygen ii. supplemental oxygen only (mask or nasal cannula); iii. continuous positive airway pressure (CPAP); iv. invasive mechanical ventilation (IMV), extra-corporeal membrane oxygenation (ECMO)). Symptoms were recorded with the Patient Symptom Questionnaire developed for the PHOSP-COVID Study;[12] responses were restricted to either cough or breathlessness for interim analyses. Percent predicted values for Forced Vital Capacity (ppFVC) and Diffusion capacity across the Lung for carbon monoxide (ppDLco) were obtained at follow-up visits and calculated on the greater of two recordings using GLI reference equations.

### 2.3 Statistical analysis

Continuous values were presented with median and interquartile range (IQR), categories were presented with proportions. Demographics and missing data were compared between the interim follow-up sample and those without early follow-up to assess any bias, chi-squared testing was performed on non-missing categories of data. The number and proportions of people with combined missing data for established indicators of ILD disease severity, namely ppDLco, ppFVC, patient symptoms, and chest x-ray (CXR), were presented as Venn diagrams. The percentage involvement of total lung damage, ground glass opacities and reticulations on CT was summarised descriptively. The lag between discharge and CT scan or research visit was presented in histograms. Bayes models were specified in order to support population estimates from small samples,[15] where prior probabilities can be iteratively updated. Analyses were performed in Stata SE16.0 within the Scottish National Safe Haven Trusted Research Environment.

To create a five-point risk strata of very low to very high risk of ILDam on CT, indicator variables and categories were dichotomised. DLco was dichotomised at 80% predicted value; FVC was dichotomised at 80% predicted value; WHO clinical progression scale was dichotomised into severity groups i-ii and iii-iv; patient symptom questionnaires were dichotomised to those who reported cough and/or breathlessness that had worsened since discharge; CXR reports were dichotomised to normal or abnormal i.e. those with a classification of “suggestive of lung fibrosis”, “extensive persistent changes greater than 1/3 lung involvement” and “indeterminate” compared with “other” or “normal”; body mass index (BMI) was dichotomised at a clinical definition of obesity (30+); IMD was dichotomised at the first two most deprived quintiles. Univariate relative risk ratios for outcomes of persistent damage >10% were estimated using Bayes binomial regression with 10000 Markov Chain Monte Carlo (MCMC) simulations and a burn-in of 10000 to support convergence. Non-informative, flat priors were selected and estimates are reported with 95% credible interval (95%CrI). Clinical indicators with significant effects were selected to develop risk strata of suspected Post-COVID ILDam.

For the indexing of risk strata in the interim cohort, missing data on indicators were imputed to the reference (lowest risk) category. Missing data within the interim follow-up sample were assumed to be not-at-random; a participant without a record of CXR, lung function tests, or cough and breathlessness recorded on the Patient Symptom Questionnaire could be less likely to have presented with these clinical indications, along with limitations on interim ascertainment by site and timing. The index, developed in those with CT scores, was applied to the remaining interim sample without scores and the percentage of participants within moderate to very-high risk strata were defined as at-risk of suspected Post-COVID ILDam.

Bayesian inference with binomial distribution of cases and non-cases, as specified by risk strata in interim data without linked CT, was used to estimate the prevalence of suspected Post-COVID ILDam within 240 days of discharge reported with the 95%CrI. MCMC simulations were run 12,500 times with a burn-in of 2,500 and random-walk Metropolis Hastings sampling. Non-informative, uniform, beta priors were used and compared in sensitivity analyses with uniform Jeffrey’s priors, as well as sceptical priors informed by published literature with or without power prior set at half the weighting. Priors of ILD prevalence were informed by population studies.[16, 17] Risk stratification and prevalence estimation was performed on the overall UKILD interim cohort, and in sensitivity analyses restricted to Tier 2 participants only.

## 3.0 Results

### 3.1 Cohort demographics and patterns of lung damage

A total of 3702 PHOSP-COVID participants reached criteria for inclusion in the interim UKILD cohort. This included, 1296 patients with data available through routine clinical care (Tier 1) and 2406 who had completed an early follow-up research visit within 240 days of discharge (Tier 2; Figure 1). We observed that 422/3702 people of the interim cohort (11.4%) had a CT scan performed, 311/2406 were performed in Tier 2 participants (12.9%) and 111/1296 were performed in Tier 1 participants (8.6%, p<0.001). A total of 87 visually scored CT scans performed within 240 days of discharge (median 119 days; IQR 83 to 155) were linked to the UKILD interim cohort (Supplementary Figure 1). Participants with a CT scored were majority male (73.6%), white (63.2%), had a median age of 58 (53 to 68) and had a median time to early follow-up visit of 149 days (IQR 121 to 171) (Table 1).

**Table 1:**
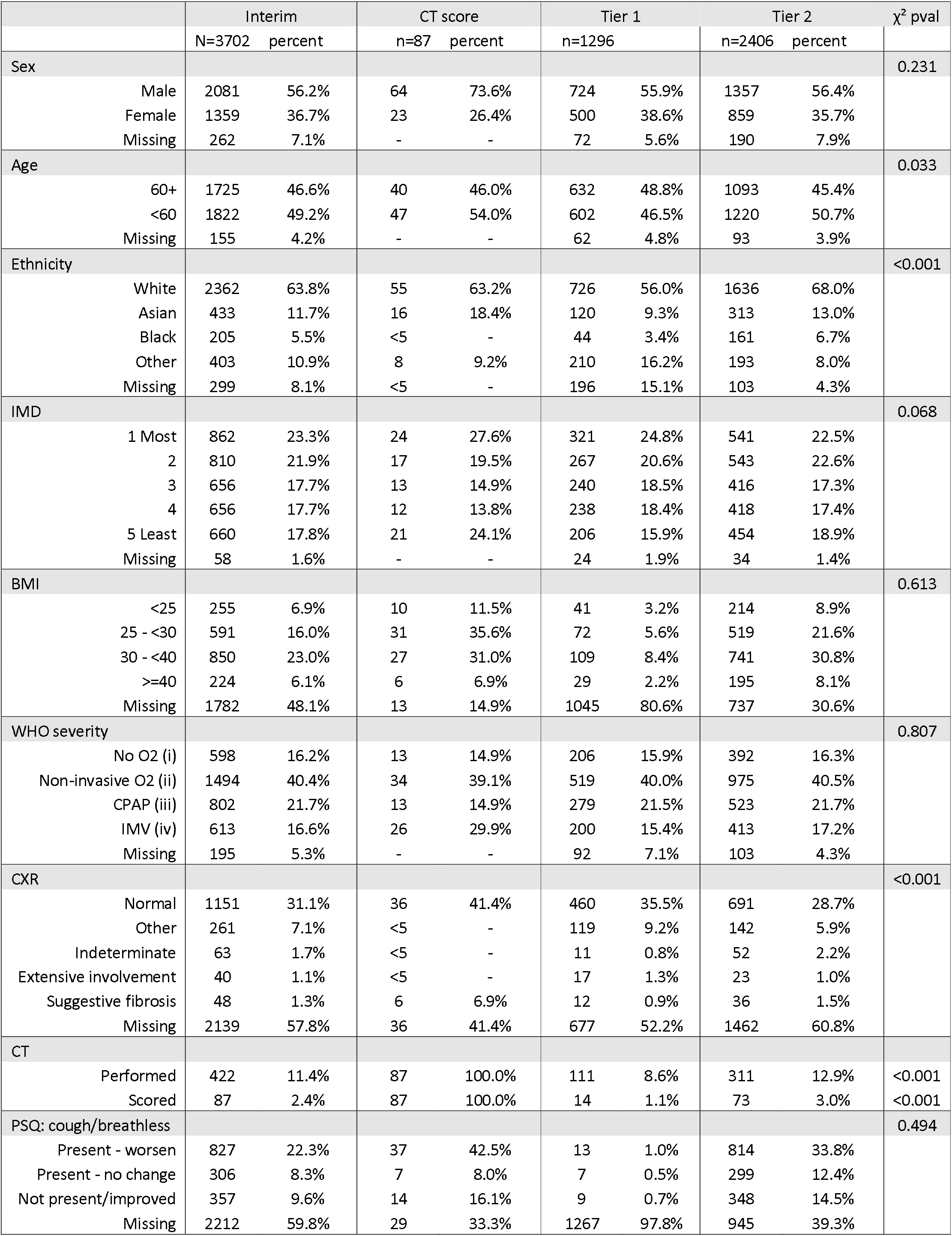
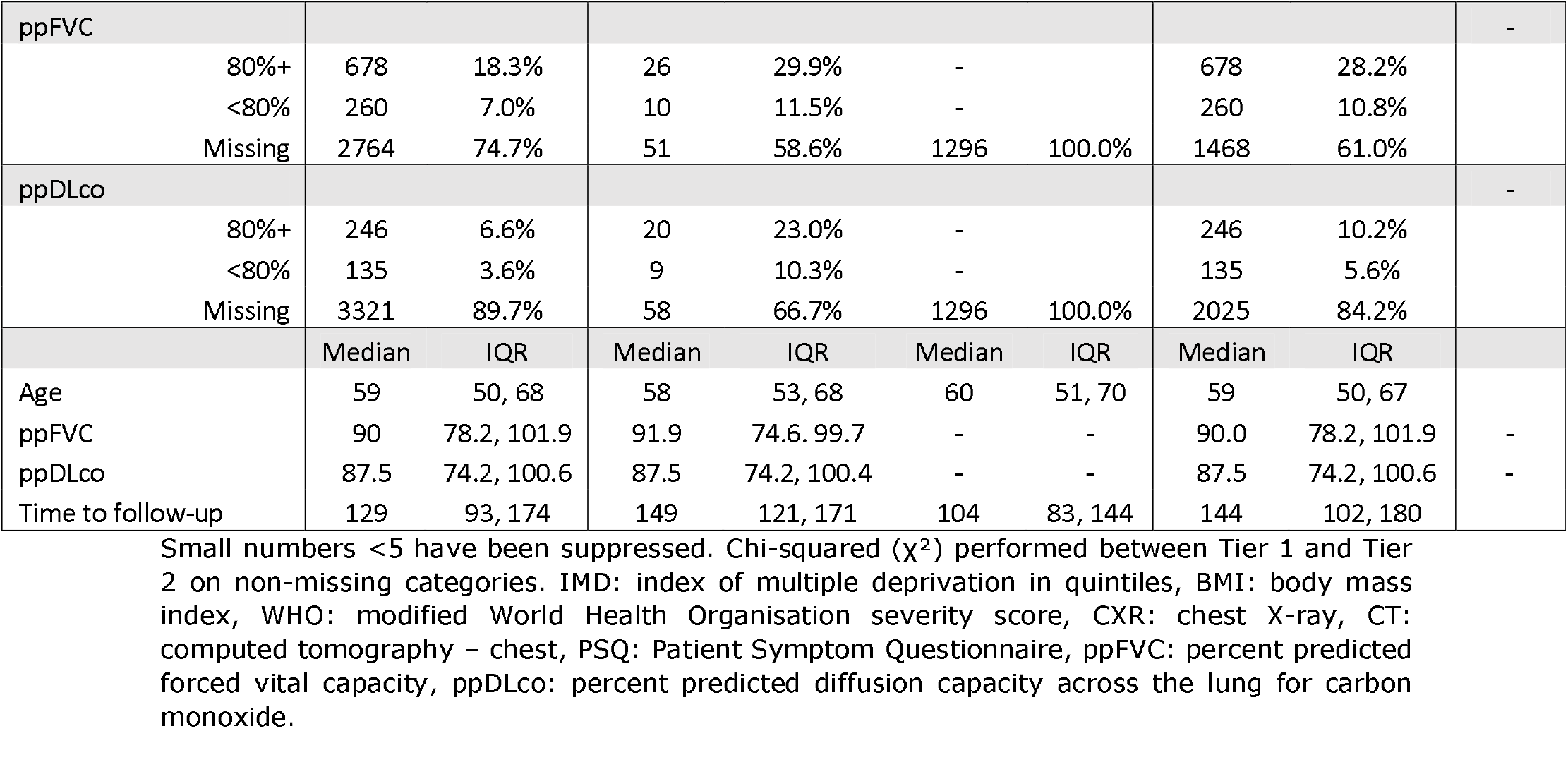
UKILD interim cohort research visit demographics.

| | | Interim | | CT score | | Tier 1 | | Tier 2 | | $\chi^2$ pval |
| --- | --- | --- | --- | --- | --- | --- | --- | --- | --- | --- |
|  |  | N=3702 | percent | n=87 | percent | n=1296 |  | n=2406 | percent |  |
| Sex |  |  |  |  |  |  |  |  |  | 0.231 |
|  | Male | 2081 | 56.2% | 64 | 73.6% | 724 | 55.9% | 1357 | 56.4% |  |
|  | Female | 1359 | 36.7% | 23 | 26.4% | 500 | 38.6% | 859 | 35.7% |  |
|  | Missing | 262 | 7.1% | - | - | 72 | 5.6% | 190 | 7.9% |  |
| Age |  |  |  |  |  |  |  |  |  | 0.033 |
|  | 60+ | 1725 | 46.6% | 40 | 46.0% | 632 | 48.8% | 1093 | 45.4% |  |
|  | <60 | 1822 | 49.2% | 47 | 54.0% | 602 | 46.5% | 1220 | 50.7% |  |
|  | Missing | 155 | 4.2% | - | - | 62 | 4.8% | 93 | 3.9% |  |
| Ethnicity |  |  |  |  |  |  |  |  |  | <0.001 |
|  | White | 2362 | 63.8% | 55 | 63.2% | 726 | 56.0% | 1636 | 68.0% |  |
|  | Asian | 433 | 11.7% | 16 | 18.4% | 120 | 9.3% | 313 | 13.0% |  |
|  | Black | 205 | 5.5% | <5 | - | 44 | 3.4% | 161 | 6.7% |  |
|  | Other | 403 | 10.9% | 8 | 9.2% | 210 | 16.2% | 193 | 8.0% |  |
|  | Missing | 299 | 8.1% | <5 | - | 196 | 15.1% | 103 | 4.3% |  |
| IMD |  |  |  |  |  |  |  |  |  | 0.068 |
|  | 1 Most | 862 | 23.3% | 24 | 27.6% | 321 | 24.8% | 541 | 22.5% |  |
|  | 2 | 810 | 21.9% | 17 | 19.5% | 267 | 20.6% | 543 | 22.6% |  |
|  | 3 | 656 | 17.7% | 13 | 14.9% | 240 | 18.5% | 416 | 17.3% |  |
|  | 4 | 656 | 17.7% | 12 | 13.8% | 238 | 18.4% | 418 | 17.4% |  |
|  | 5 Least | 660 | 17.8% | 21 | 24.1% | 206 | 15.9% | 454 | 18.9% |  |
|  | Missing | 58 | 1.6% | - | - | 24 | 1.9% | 34 | 1.4% |  |
| BMI |  |  |  |  |  |  |  |  |  | 0.613 |
|  | <25 | 255 | 6.9% | 10 | 11.5% | 41 | 3.2% | 214 | 8.9% |  |
|  | 25 - <30 | 591 | 16.0% | 31 | 35.6% | 72 | 5.6% | 519 | 21.6% |  |
|  | 30 - <40 | 850 | 23.0% | 27 | 31.0% | 109 | 8.4% | 741 | 30.8% |  |
|  | >=40 | 224 | 6.1% | 6 | 6.9% | 29 | 2.2% | 195 | 8.1% |  |
|  | Missing | 1782 | 48.1% | 13 | 14.9% | 1045 | 80.6% | 737 | 30.6% |  |
| WHO severity |  |  |  |  |  |  |  |  |  | 0.807 |
|  | No O2 (i) | 598 | 16.2% | 13 | 14.9% | 206 | 15.9% | 392 | 16.3% |  |
|  | Non-invasive O2 (ii) | 1494 | 40.4% | 34 | 39.1% | 519 | 40.0% | 975 | 40.5% |  |
|  | CPAP (iii) | 802 | 21.7% | 13 | 14.9% | 279 | 21.5% | 523 | 21.7% |  |
|  | IMV (iv) | 613 | 16.6% | 26 | 29.9% | 200 | 15.4% | 413 | 17.2% |  |
|  | Missing | 195 | 5.3% | - | - | 92 | 7.1% | 103 | 4.3% |  |
| CXR |  |  |  |  |  |  |  |  |  | <0.001 |
|  | Normal | 1151 | 31.1% | 36 | 41.4% | 460 | 35.5% | 691 | 28.7% |  |
|  | Other | 261 | 7.1% | <5 | - | 119 | 9.2% | 142 | 5.9% |  |
|  | Indeterminate | 63 | 1.7% | <5 | - | 11 | 0.8% | 52 | 2.2% |  |
|  | Extensive involvement | 40 | 1.1% | <5 | - | 17 | 1.3% | 23 | 1.0% |  |
|  | Suggestive fibrosis | 48 | 1.3% | 6 | 6.9% | 12 | 0.9% | 36 | 1.5% |  |
|  | Missing | 2139 | 57.8% | 36 | 41.4% | 677 | 52.2% | 1462 | 60.8% |  |
| CT |  |  |  |  |  |  |  |  |  |  |
|  | Performed | 422 | 11.4% | 87 | 100.0% | 111 | 8.6% | 311 | 12.9% | <0.001 |
|  | Scored | 87 | 2.4% | 87 | 100.0% | 14 | 1.1% | 73 | 3.0% | <0.001 |
| PSQ: cough/breathless |  |  |  |  |  |  |  |  |  | 0.494 |
|  | Present - worsen | 827 | 22.3% | 37 | 42.5% | 13 | 1.0% | 814 | 33.8% |  |
|  | Present - no change | 306 | 8.3% | 7 | 8.0% | 7 | 0.5% | 299 | 12.4% |  |
|  | Not present/improved | 357 | 9.6% | 14 | 16.1% | 9 | 0.7% | 348 | 14.5% |  |
|  | Missing | 2212 | 59.8% | 29 | 33.3% | 1267 | 97.8% | 945 | 39.3% |  |

|  |  |  |  |  |  |  |  |  |  |
| --- | --- | --- | --- | --- | --- | --- | --- | --- | --- |
| ppFVC |  |  |  |  |  |  |  |  | - |
| 80%+ | 678 | 18.3% | 26 | 29.9% | - |  | 678 | 28.2% |  |
| <80% | 260 | 7.0% | 10 | 11.5% | - |  | 260 | 10.8% |  |
| Missing | 2764 | 74.7% | 51 | 58.6% | 1296 | 100.0% | 1468 | 61.0% |  |
| ppDLco |  |  |  |  |  |  |  |  | - |
| 80%+ | 246 | 6.6% | 20 | 23.0% | - |  | 246 | 10.2% |  |
| <80% | 135 | 3.6% | 9 | 10.3% | - |  | 135 | 5.6% |  |
| Missing | 3321 | 89.7% | 58 | 66.7% | 1296 | 100.0% | 2025 | 84.2% |  |
|  | Median | IQR | Median | IQR | Median | IQR | Median | IQR |  |
| Age | 59 | 50, 68 | 58 | 53, 68 | 60 | 51, 70 | 59 | 50, 67 |  |
| ppFVC | 90 | 78.2, 101.9 | 91.9 | 74.6, 99.7 | - | - | 90.0 | 78.2, 101.9 | - |
| ppDLco | 87.5 | 74.2, 100.6 | 87.5 | 74.2, 100.4 | - | - | 87.5 | 74.2, 100.6 | - |
| Time to follow-up | 129 | 93, 174 | 149 | 121, 171 | 104 | 83, 144 | 144 | 102, 180 |  |

Small numbers <5 have been suppressed. Chi-squared ( $\chi^2$ ) performed between Tier 1 and Tier 2 on non-missing categories. IMD: index of multiple deprivation in quintiles, BMI: body mass index, WHO: modified World Health Organisation severity score, CXR: chest X-ray, CT: computed tomography – chest, PSQ: Patient Symptom Questionnaire, ppFVC: percent predicted forced vital capacity, ppDLco: percent predicted diffusion capacity across the lung for carbon monoxide.

**Figure 1.**
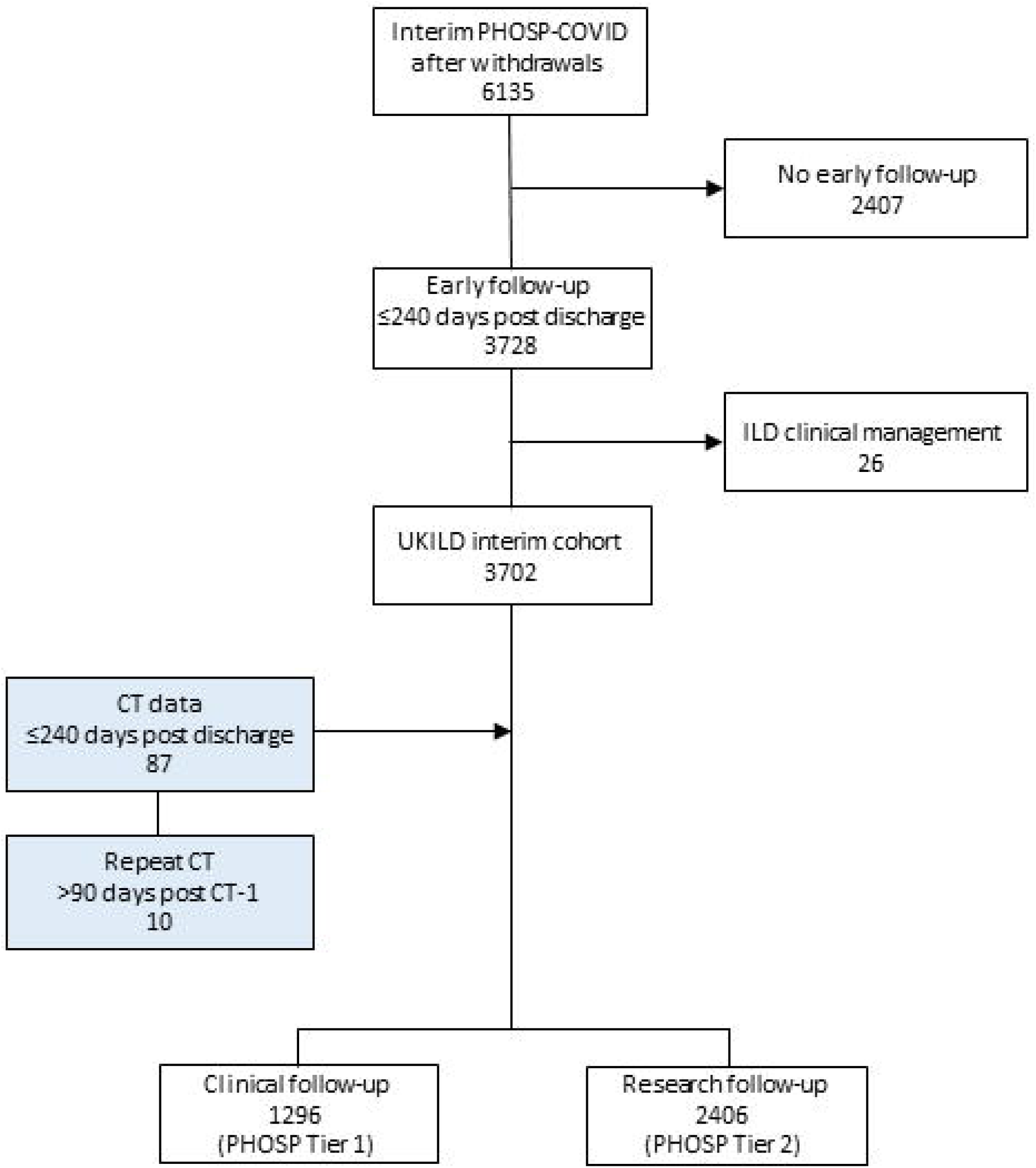
Flow diagram of UKILD interim cohort definition. White boxes derived from PHOSP-COVID database. Blue boxes represent CT scored sample identified through PHOSP-COVID to a linked radiological database.

ILDam was visually observed in 74/87 scans (85.1%). Visual scoring of the interstitial lung damage revealed ground glass opacities affecting a median 20.8% (IQR 10.8 to 30.0) of the lung, and reticulation at a median 15.0% (IQR 7.5 to 22.5) lung involvement, with a median total interstitial lung damage of 36.7% (IQR 22.3 to 51.7) (Figure 2A, Supplementary Table 1). 10 people had a repeat CT visually scored after a minimum of 90 days (median 168 days; IQR 166 to 213), 9 of whom were classified with ILDam on the initial scan, whilst one person no longer reached the threshold. In 8 participants, comparable levels of lung involvement of ground glass opacities, reticulations and total lung damage were observed between scans (Figure 2B-D, Supplementary Table 1).

**Figure 2.**
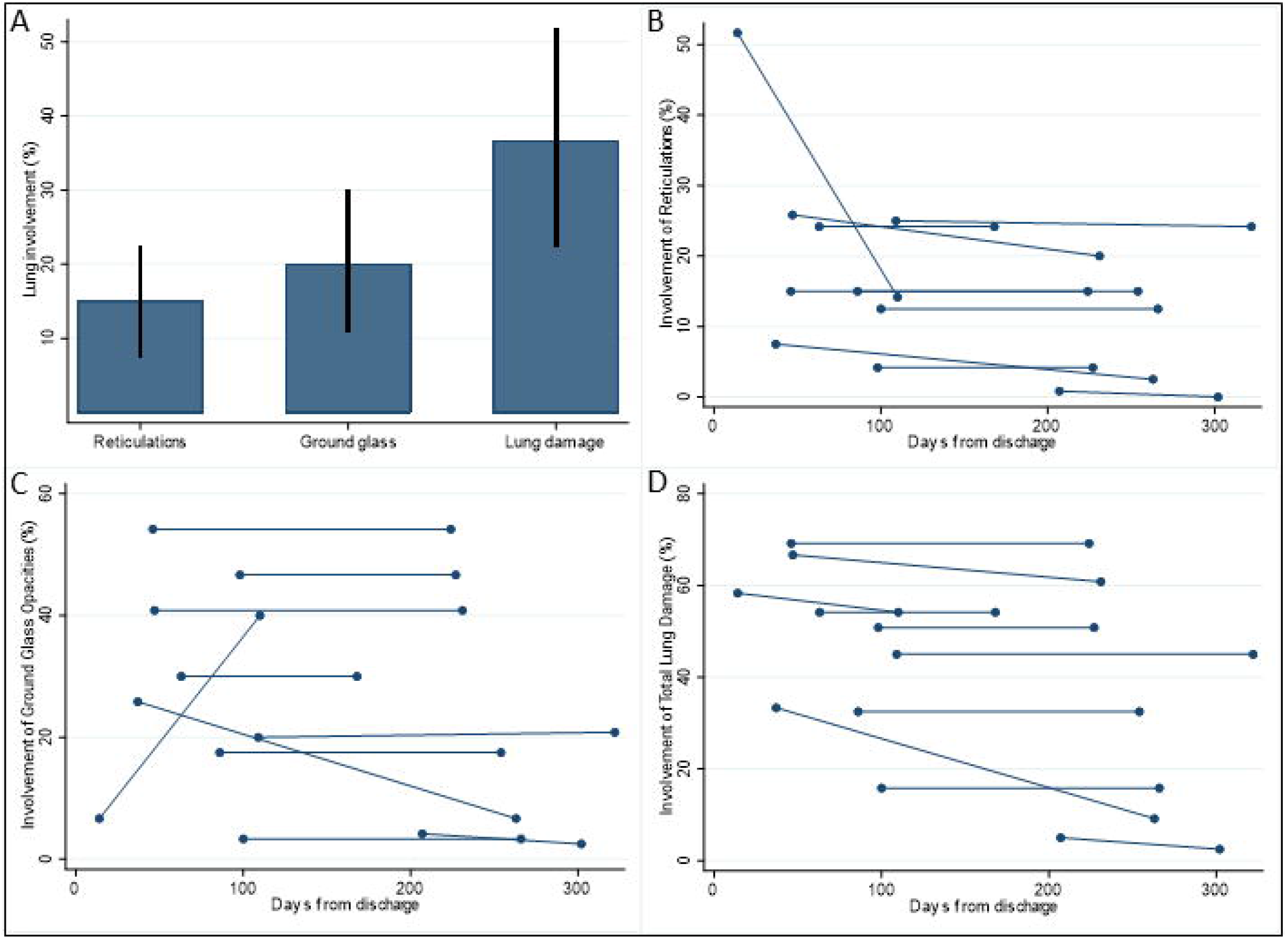
Visually scored radiological patterns on linked CT. Percentage lung involvement of reticulations, ground glass opacities, and total lung damage in A) CT scans within 240 days of discharge with visually scored persistent damage >10% involvement (n=74). Percentage lung involvement of B) reticulations, C) ground glass opacities and D) total lung damage at initial and repeat CT scans with >90days between (n=10). Box plots present median and IQR, line plots present individual involvement across scan 1 and scan 2 with days from hospital discharge presented on the x-axis.

Overall, the median time to follow-up in the UKILD interim cohort (N=3702) was 129 days (IQR 83 to 174), the median age was 59 (IQR 50 to 68) and the cohort was majority male (56.2%). Tier 1 participants (n=1296) had a median time to follow-up of 104 days (IQR 83 to 144), a median age of 60 (IQR 51 to 70) and the majority were male (55.9%); demographics were similar in Tier 2 participants (n=2406) with a median time to research visit of 144 days (IQR 102 to 180), a median age of 59 (IQR 50 to 67) and a majority male (56.1%) (Table 1). There was minimal evidence of systematic bias in the characteristics between Tier 2 and Tier 1 participants in non-missing data (Table 1), although the representation of people aged below 60 was greater in Tier 2 participants (50.7% vs 46.5%; p=0.033), similarly there was also more representation from people with white background (68.0% vs 56.0%; p<0.001), as well as lower representation of normal CXR (28.7% vs 35.5%; p<0.001). Tier 2 participants had a median ppFVC of 90.0% (IQR 78.2 to 101.9) with missing records at 61.0%, whilst median ppDLCO was 87.5% (IQR 74.2 to 100.6) with missing records at 84.2%; lung function was missing in follow-up of Tier 1 participants. We observed 33.8% of people reported worsening cough or dyspnoea since discharge in Tier 2.

Missing data were frequently substantial in the UKILD interim cohort across both Tier 1 and Tier 2, including BMI (48.1%), CXR (57.8%), and Patient Symptom Questionnaires (cough and/or dyspnoea, 59.8%). Missing data across four diagnostic indicators of ILD (FVCpp, TLCOpp, CXR, and Patient Symptom Questionnaires) were described separately in each Tier (Figure 3). In Tier 1, 661/1296 (51.0%) were missing data on all four characteristics at interim, whilst in Tier 2 473/2406 people (19.7%) were missing data on all four characteristics. In contrast, a total of only 149 Tier 2 participants had complete data on all (6.2%), whilst no Tier 1 participants had complete data on all four indicators. Missing data were also frequent when restricted to people with visual scores of interstitial lung damage (n=87), particularly for lung function (ppDLco 66.7%; ppFVC 58.6%), CXR (41.4%), and Patient Symptom Questionnaire (33.3%) (Table 1).

**Figure 3.**
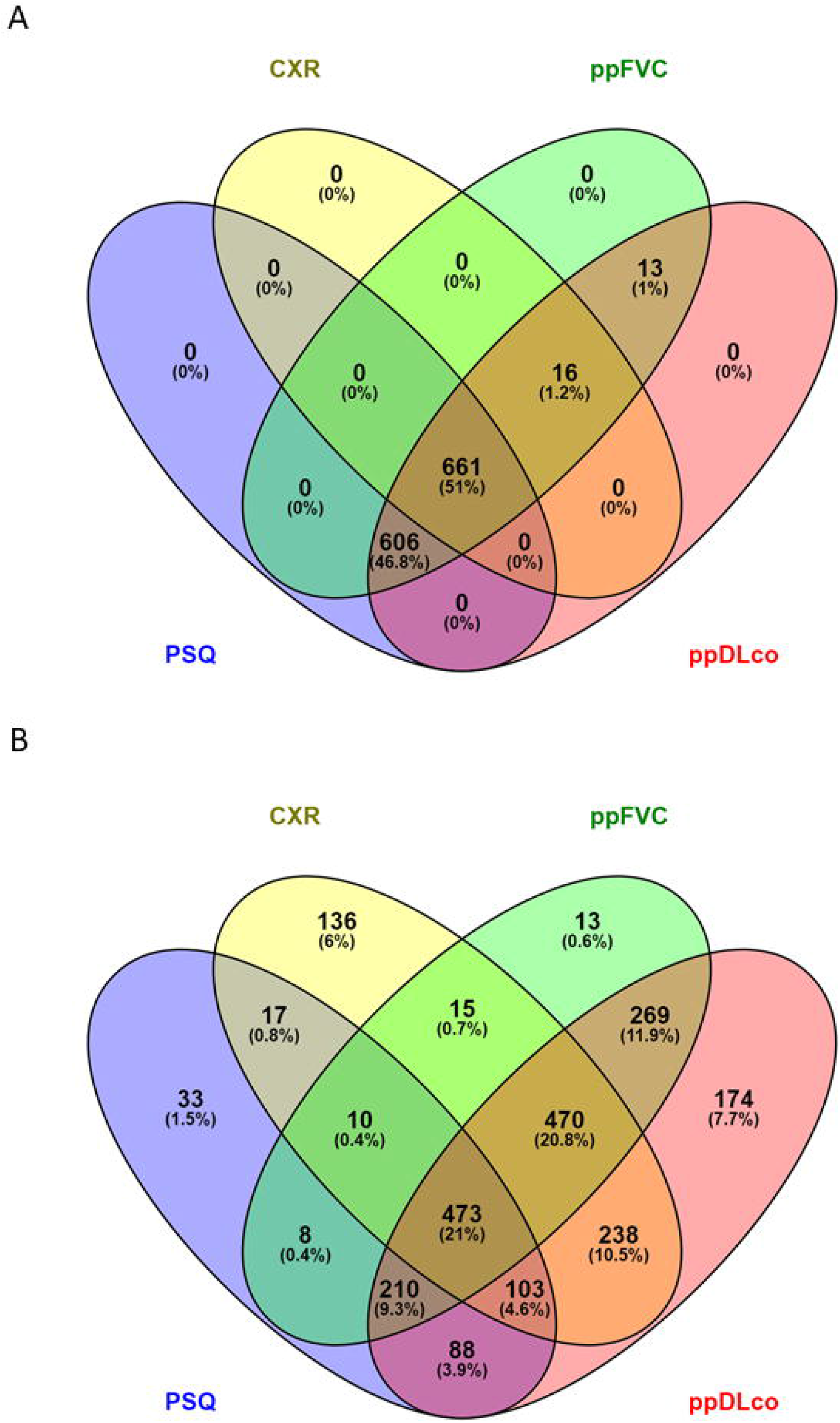
Missing records in ILD diagnostic indicators. Missing data and percentage are reported for Tier 1 (n=1296) and Tier 2 (n=2406) according to ppFVC (percent predicted forced vital capacity), ppDLCO (percent predicted DLco gas transfer), PSQ (patient symptom questionnaire, cough and/or breathlessness), CXR (chest X-ray). *Venny (2007-2015) https://bioinfogp.cnb.csic.es/tools/venny/index.html*

### 3.2 Risk of interstitial lung damage and suspected Post-COVID ILDam

Univariate risk ratios were calculated to assess the risk of visually scored ILDam >10% on CT. A greater risk of ILDam was observed in males (RR 1.48 95%CrI 1.13; 2.09) and in those over 60 years of age (RR 1.17 95%CrI 1.00; 1.44). Clinical indicators, including severe illness on admission requiring CPAP, IMV or ECMO (RR 1.27 95%CrI 1.07; 1.56), abnormal CXR findings (RR 1.21 95%CrI 1.05; 1.40), and ppDLco <80% (RR 1.25 95%CrI 1.00; 1.56) were associated with greater risk (Table 2).

**Table 2:** Univariate risk ratio of visually scored interstitial lung damage.

| Characteristic | ILDam>10% (n) | Percent of non-missing cases | Univariate risk ratio | 95% Credible Interval |
| --- | --- | --- | --- | --- |
| Male | 59 | 79.7% | 1.48 | 1.13; 2.09 |
| Age 60+ | 37 | 50.0% | 1.17 | 1.00; 1.44 |
| Non-white | 24 | 33.8% | 0.83 | 0.63; 1.05 |
| IMD (Q1/2) | 38 | 51.4% | 1.19 | 0.99; 1.43 |
| BMI >30 | 31 | 49.2% | 1.20 | 0.99; 1.48 |
| CPAP/IMV | 38 | 51.4% | 1.27 | 1.07; 1.55 |
| aCXR | 11 | 25.0% | 1.21 | 1.05; 1.40 |
| ppFVC <80 | 9 | 28.1% | 1.01 | 0.79; 1.46 |
| ppDLco <80 | 9 | 36.0% | 1.25 | 1.00; 1.56 |
| PSQ worse | 32 | 62.7% | 0.99 | 0.73; 1.17 |

Percentage of exposed cases from non-missing case observations presented with risk ratio (RR) and 95% credible interval derived from binomial regression. Interstitial lung damage (ILDam); Body mass index (BMI); continuous positive airway pressure or invasive mechanical ventilation (CPAP/IMV); abnormal chest x-ray (aCXR); percent predicted forced vital capacity (ppFVC); percent predicted diffusion capacity across the lung for carbon monoxide (ppDLco); Patient Symptom Questionnaire (PSQ).

Three significant clinical indicators were selected to index the risk of Post-COVID ILDam in the overall cohort based on combined thresholds: ppDLco <80%; abnormal CXR; and severe illness on admission. Missing data on dichotomised thresholds were imputed at the reference category for these indicators. We considered individuals to be at very-high risk when reaching the defined thresholds in all three indicators (risk index 4), high risk when two thresholds were reached (risk index 3), or moderate risk if reaching ppDLco or CXR thresholds alone (risk index 2). Individuals reaching the threshold of severity of illness on admission alone were considered low-risk in the absence of other indicators (risk index 1). Those who did not reach any threshold were considered very low risk (risk index 0). In participants who had either received a CT that had not been scored (n=335), or not received a CT (n=3280), a total 12/3615 participants (0.3%) were considered very-high risk, 133/3615 at high risk (3.7%), and 104/3615 at moderate risk (2.9%), 1236/3615 at low risk (34.2%) and 2130/3615 at very-low risk (58.9%) (Table 3). Combined, 249/3615 (6.9%) people in strata of moderate to very-high risk were defined as at risk of Post-COVID ILDam in those without CT scores. In sensitivity analyses applying risk stratification to Tier 2 alone, 210/2333 (9.0%) of participants were at moderate to very-high risk of Post-COVID ILDam (Table 3).

**Table 3:** Suspected Post-COVID ILDam risk strata in interim UKILD cohort.

| Risk<br>(UKILD interim) | No CT<br>(3280) | CT unscored<br>(335) | PC-ILD risk<br>(3615) | Percent |
| --- | --- | --- | --- | --- |
| Very high | 6 | 6 | 12 | 0.3% |
| High | 92 | 41 | 113 | 3.7% |
| Moderate | 84 | 20 | 104 | 2.9% |
| Low | 1119 | 117 | 1236 | 34.2% |
| Very low | 1979 | 151 | 2130 | 58.9% |
| Risk<br>(Tier 2 Sensitivity) | No CT<br>(2095) | CT unscored<br>(238) | PC-ILD risk<br>(2333) | Percent |
| Very high | 6 | 6 | 12 | 0.5% |
| High | 83 | 35 | 118 | 5.1% |
| Moderate | 64 | 16 | 80 | 3.4% |
| Low | 707 | 71 | 778 | 33.3% |
| Very low | 1235 | 110 | 1345 | 57.7% |

Risk strata: very high – all three ILD diagnostic indicators present (abnormal CXR, ppDLco <80%, severe admission requiring CPAP or IMV). High – at least two indicators present. Moderate – either abnormal CXR or ppDLco<80% present. Low – severe admission present. Very low – no indicators present. Missing data were imputed at the reference category. Percent denominator is interim cohort without linked, scored CT (n=3615; Tier 2 sensitivity n=2333).

The demographics of the at-risk group (n=249) were compared to those with visually scored ILDam (n=74) (Supplementary Table 2). No differences were observed according to representation of males, older age, ethnicity, IMD, BMI, severity of admission, ppFVC <80% or Patient Symptom Questionnaire. There was lower representation of normal CXR in the at-risk group compared to those with visually scored ILDam (11.6% vs 39.2%, p<0.001) and more representation of ppDLco <80% (50.6% vs 12.2%, p<0.001). We observed that CXR was missing in 26.5% of people in the at-risk group, whilst 26.9% of these individuals had a CT performed.

Based on the distribution of Post-COVID ILDam risk cases defined in the UKILD interim cohort above, the prevalence of suspected Post-COVID ILDam up to 240 days after hospitalisation was estimated between 6.5% and 8.3% with 95% credibility (7.42%, 95%CrI 6.55; 8.34) using non-informative priors. This estimate reduced to between 5.0% and 6.4% (5.71%, 95%CrI 5.03; 6.41) with sceptical priors based on ILD population prevalence estimated at 1 in 1,000 (Supplementary Table 3, Supplementary Figure 2).[16, 17] In sensitivity analyses restricted to Tier 2 cases, the prevalence of suspected Post-COVID ILDam was estimated between 8.7% and 11.3% with 95% credibility (9.92%, 95%CrI 8.69; 11.25) using non-informative priors, or between 5.9% and 7.7% using sceptical priors (6.73%, 95%CrI 5.85; 7.67).

## 4.0 Discussion

These data demonstrate that interstitial lung damage (ILDam) was visually identifiable on clinically indicated follow-up CT imaging before eight months in a substantial proportion of patients discharged from hospital following COVID-19, and demonstrated minimal resolution in subsequent scans where follow-up scans were available. Key clinical indicators including abnormal CXR, ppDLco <80% and severe admissions requiring invasive support (IMV, CPAP, ECMO) were associated with ILDam in the eight month period after discharge. We estimate the percentage of people at very-high risk of Post-COVID ILDam as 0.3% (all three indicators present), high risk as 3.7% (any two indicators present), and moderate risk as 2.9% (presence of either ppDLco<80% or abnormal CXR, alone). Combining these risk strata, 6.9% of the interim cohort had suspected Post-COVID ILDam, which increased to 9.0% in sensitivity analysis on those with research follow-up visits. Based on modelling with non-informative as well as sceptical priors, in both overall and sensitivity analyses, we estimate the prevalence of suspected Post-COVID ILDam to be between 5.0% and 11.3% up to 240 days post-hospitalisation, for acute COVID-19 infections before March 2021.

It is not possible to determine, at the time of this interim analysis, whether the observed ILDam represents early Interstitial Lung *DISEASE* (ILD) with potential for progression, or whether it reflect residual pneumonitis or post-Acute Respiratory Distress Syndrome (ARDS) which are known to be stable or resolve over time.[18] Hence for these analyses we refer to interstitial lung *DAMAGE (ILDam)* as we recognise further follow-up and mechanistic studies will be required to determine the clinical trajectory of these observations. Where linked longitudinal scans were available most patients did not show evidence of improvement, although such clinically requested CTs may be over-represented by those with slower recovery. However, nearly half the people with visually scored ILDam did not require CPAP, IMV or ECMO during their admission, suggesting that in the medium and longer term there may be considerable disability in those suffering from Post-COVID ILDam consistent with prior studies. [18]

Primary analyses of the UKILD-Post COVID ILD study will use complete data from hospitalised and non-hospitalised participants to provide updated estimates of Post-COVID ILD at both early and late follow-up, where quantitation of airway disease parameters on radiological imaging is ongoing.[13] The extent of missing data for ILD diagnostic indicators was high at interim analysis, despite this, 6.9% of PHOSP-COVID participants were at risk of Post-COVID ILDam, which requires both CT confirmation and lung function follow-up that were frequently incomplete. Restricting analyses to Tier 2, with less missing data, suggested 9.0% of PHOSP-COVID participants were at risk. CT scans were only performed when clinically indicated and healthcare service prioritisation changed during the COVID-19 pandemic. Similarly, lung function testing, especially measurement of DLco, was severely restricted in many sectors due to enhanced infection control procedures during the pandemic. However, considering approximately 459,000 people were hospitalised with COVID-19 in the UK National Health Service by end of March 2021,[19] our interim findings suggest Post-COVID ILDam is not uncommon and should emphasise the importance of active radiological and physiological monitoring especially in people at moderate, or above, risk of ILDam.[14]

### 4.1 Strengths and Limitations

This interim analysis of the UKILD Post-COVID study is the largest assessment of ILDam in patients hospitalised for COVID-19 to date, and the findings are consistent with findings from a number of smaller studies that demonstrate persistent radiological patterns and impaired gas transfer during extended follow-up of patients with COVID-19.[20-24] For participants in receipt of a clinically indicated but unscored CT, we observed that 67/335 (20.0%) people were in moderate to very-high risk strata (sensitivity 57/238, 24.0%), which was similar to the percentage of CT scans with radiological patterns suggestive of fibrosis within the first year post-hospitalisation estimated in meta-analysis (29%; 95%CI 22% to 37%).[25] The UKILD long-COVID cohort excluded participants with any evidence of ILD prior to hospitalisation, and we used informative sceptical priors and power priors for more conservative estimates, which continued to suggest a substantial burden of Post-COVID ILDam. The approach we report can be reasonably applied to later follow-up, with current findings used as informative priors for updating Bayesian inference.

Whilst included CTs were assumed to be representative of clinically indicated radiology, this is limited by local management protocols and timing of services, which increases chances of selection and ascertainment bias. Furthermore, individuals with linked CT may have unrecorded pre-existing disease or present with radiological patterns suggestive of emphysema or end-stage COPD, which required disentangling from COVID-19 sequelae. CT scoring was performed pragmatically to estimate residual burden of COVID-19 disease. Visual scores of ILDam >10% involvement was defined here as suspected Post-COVID ILDam.

We recognise these interim findings may also be limited by misclassification. Descriptive analyses identified substantial missing data in clinical indicators of ILDam, limiting imputation and multivariable modelling. We used dichotomised thresholds with missing data imputed at the reference category to support risk strata classification and maintain denominators. It is plausible that participants who reached the risk thresholds clinically may have been otherwise categorised in the absence of missing data records. Similarly, lung involvement of reticulation and ground glass opacities was frequently observed in clinically indicated CTs and some individuals were scored with ILDam on CT who did not reach the moderate risk strata due to missing indicator variables. These limitations may underestimate the number at moderate to very high risk at early follow-up. The PHOSP-COVID Study data collection is ongoing, missing data in further UKILD analyses are likely to be fewer as more data are inputted and linked to clinical data following interim data freeze. Furthermore, missing data are likely to be at random, for which multiple imputation approaches will be appropriate.

Finally we recognise that these findings may not be generalisable to all populations especially people not admitted to hospital. Severe admissions requiring CPAP or IMV were over-represented in the PHOSP-COVID dataset relative to hospitalised survivors of COVID-19,[12] which may also inflate prevalence estimates as we were unable to classify ARDS events. Furthermore, these data reflect people who were discharged before end of March 2021, and do not represent later SARS-CoV-2 variants in fully vaccinated populations that more frequently led to milder infections.

### 4.2 Conclusion

Whilst missing data in observational cohort studies is a severe limitation, we demonstrate that thresholds of ppDLco, CXR and severity of admission can stratify risk of ILDam involving more than 10% of the lung. In a limited sample size, similar ILDam was observed in subsequent scans, suggesting minimal resolution although the functional consequence is currently unknown. These findings highlight the importance of radiological and physiological monitoring of patients at both early and later follow-up, particularly for individuals meeting criteria for moderate risk strata for Post-COVID ILDam. These interim data will be used to inform clinical management and research strategies to elucidate the development and functional implication of Post-COVID ILDam.

## Data Availability

Data were obtained through the PHOSP-COVID Study

https://www.phosp.org/

## Acknowledgements

Jointly funded by UK Research and Innovation and National Institute of Health Research (grant references: MR/V027859/1 and COV0319). Ethics Approval Ethics Ref: 20/YH/0225

The authors would like to acknowledge the support of the eDRIS Team (Public Health Scotland) for their involvement in obtaining approvals, provisioning and linking data and the use of the secure analytical platform within the National Safe Haven.

This study would not be possible without all the participants who have given their time and support. We thank all the participants and their families. We thank the many research administrators, health-care and social-care professionals who contributed to setting up and delivering the study at all of the 65 NHS trusts/Health boards and 25 research institutions across the UK, as well as all the supporting staff at the NIHR Clinical Research Network, Health Research Authority, Research Ethics Committee, Department of Health and Social Care, Public Health Scotland, and UK Health Security Agency, and support from the ISARIC Coronavirus Clinical Characterisation Consortium (ISARIC4C). We thank Kate Holmes at the NIHR Office for Clinical Research Infrastructure (NOCRI) for her support in coordinating the charities group.

The PHOSP-COVID industry framework was formed to provide advice and support in commercial discussions, and we thank the Association of the British Pharmaceutical Industry as well as Ivana Poparic and Peter Sargent at NOCRI for coordinating this. We are very grateful to all the charities that have provided insight to the study: Action Pulmonary Fibrosis, Alzheimer’s Research UK, Asthma & Lung UK, British Heart Foundation, Diabetes UK, Cystic Fibrosis Trust, Kidney Research UK, MQ Mental Health, Muscular Dystrophy UK, Stroke Association Blood Cancer UK, McPin Foundations, and Versus Arthritis. We thank the NIHR Leicester Biomedical Research Centre patient and public involvement group and the Long Covid Support Group.

JB acknowledges MRC Transition Fellowship (MR/T032529/1) and Manchester BRC funding. BG acknowledges UKRI-MRC Programme Grant and Confidence in Concept Grant, British Lung Foundation and the NIHR Leicester BRC funding. BGG acknowledges funding from Wellcome Trust grant 221680/Z/20/Z. NG is funded by an NIHR fellowship. PLM and RJA are funded by the Action for Pulmonary Fibrosis Mike Bray Fellowships. DGW is funded by an NIHR Advanced Fellowship. JP is supported by UKRI PC-ILD grant, Breathing Matters Charity, and UCLH/UCL funding from the Department of Health’s NIHR Biomedical Research Centres funding scheme. AART is funded by an Intermediate Clinical Fellowship from the British Heart Foundation (FS/18/13/33281). LVW is supported by GSK / Asthma + Lung UK Chair in Respiratory Research (C17-1). GJ acknowledges funding from a NIHR Research Professorship. IS fellowship is funded by the Rayne Foundation.

## Author Contributions

Conceptualisation: IS, JJ, JMW, JCP, PLM, PMG, RJA, JKB, SLB, PB, SMB, JFB, NC, GC, EKD, AD, LF, MAG, FVG, BG, IPH, NAH, MH, TEH, SRJ, MGJ, FK, RL, PM, MP, KP, JKQ, PRO, MS, AJS, DJFS, MS, LGS, SS, DRT, AART, SLFW, NDW, MEW, DGW, CEB, RCC, LPH, KPH, LVW, RGJ. Data curation: EMH, MS, OL, HM, IS, JJ, NL, AB, JKQ. Formal analysis: IS, JJ, JKQ. Funding acquisition: CEB, LVW, RAE, JC, LPH, AH, MM, KP, BR, OL, MR, OE, HM, ASh, MS, RS, VH, LH, NG, RGJ. Project administration: ASh, MW, AS. Writing - original draft: IS, JJ, PMG, RGJ. Writing - review & editing: IS, JJ, PMG, PLM, JCP, RJA, JKB, SLB, PB, SMB, JFB, JC, RCC, NC, CC, GC, EKD, AD, OE, RAE, LF, MAG, FVG, BG, NJG, BGG, IPH, NAH, VH, EMH, MH, TEH, AH, LHW, IJ, SRJ, MGJ, FK, RL, OL, NL, MM, HM, PM, EO, DP, KPH, MP, JP, KP, JKQ, BR, MR, PRO, LS, RS, MGS, MS, ASh, AJS, AS, DJFS, MS, LGS, SS, DT, AART, MT, SLFW, SW, NDW, JMW, DGW, CEB, LPH, LVW, RGJ.

