## Supplementary for "Interstitial lung damage following COVID-19 hospitalisation: an interim analysis of the UKILD Post-COVID study"

**Supplemental Tables and Figures**

**Supplementary Table 1: Radiological patterns in visually scored CT scans**

|  | n | Median | IQR |
| --- | --- | --- | --- |
| Visually scored interstitial lung damage (>10%) | |  |  |
| reticulation | 74 | 15.0% | (7.5, 22.5) |
| Ground glass opacities | 74 | 20.8% | (10.8, 30.0) |
| Total damage | 74 | 36.7% | (22.3, 51.7) |
| Visually scored interstitial lung damage (<10%) | | |  |
| reticulation | 13 | 0.0% | (0.0, 0.8) |
| Ground glass opacities | 13 | 0.8% | (0.0, 3.3) |
| Total damage | 13 | 2.5% | (0.0, 4.2) |
| Repeat CT Scan Subgroup Scan: 1 | | |  |
| reticulation | 10 | 15.0% | (7.5, 25.0) |
| Ground glass opacities | 10 | 22.9% | (6.7, 40.8) |
| Total damage | 10 | 47.9% | (32.5, 58.3) |
| Repeat CT Scan Subgroup: Scan 2 | | |  |
| reticulation | 10 | 14.6% | (4.2, 20.0) |
| Ground glass opacities | 10 | 25.4% | (6.7, 40.8) |
| Total damage | 10 | 47.9% | (15.8, 54.2) |

**Supplementary Table 2: Comparison of demographics between visually scored ILDam sample and Post-COVID ILDam at-risk group**

|  | Interim | | At-risk | | ILDam | | χ² pval |
| --- | --- | --- | --- | --- | --- | --- | --- |
|  | N=3702 | percent | N=249 | percent | N=74 | percent |  |
| Sex |  |  |  |  |  |  | 0.168 |
| Male | 2081 | 56.6% | 177 | 71.1% | 59 | 79.7% |  |
| Female | 1359 | 35.6% | 70 | 28.1% | 15 | 20.3% |  |
| Age |  |  |  |  |  |  | 0.083 |
| 60+ | 1725 | 45.5% | 152 | 61.0% | 37 | 50.0% |  |
| <60 | 1822 | 50.7% | 96 | 38.6% | 37 | 50.0% |  |
| Ethnicity |  |  |  |  |  |  | 0.854 |
| White | 2362 | 67.9% | 165 | 66.3% | 47 | 63.5% |  |
| Asian | 433 | 13.0% | 35 | 14.1% | 12 | 16.2% |  |
| Black | 205 | 6.6% | 17 | 6.8% | <5 | - |  |
| Other | 403 | 8.1% | 21 | 8.4% | 8 | 10.8% |  |
| Missing | 299 | 4.4% | 11 | 4.4% | <5 | - |  |
| IMD |  |  |  |  |  |  | 0.381 |
| 1 Most | 862 | 22.5% | 57 | 22.9% | 22 | 29.7% |  |
| 2 | 810 | 22.5% | 60 | 24.1% | 16 | 21.6% |  |
| 3 | 656 | 17.3% | 43 | 17.3% | 10 | 13.5% |  |
| 4 | 656 | 17.3% | 52 | 20.9% | 11 | 14.9% |  |
| 5 Least | 660 | 18.9% | 35 | 14.1% | 15 | 20.3% |  |
| BMI |  |  |  |  |  |  | 0.458 |
| <25 | 255 | 8.9% | 28 | 11.2% | 8 | 10.8% |  |
| 25 - <30 | 591 | 21.5% | 77 | 30.9% | 24 | 32.4% |  |
| 30 - <40 | 850 | 30.8% | 81 | 32.5% | 26 | 35.1% |  |
| >=40 | 224 | 8.1% | 16 | 6.4% | 5 | 6.8% |  |
| Missing | 1782 | 30.7% | 47 | 18.9% | 11 | 14.9% |  |
| WHO severity |  |  |  |  |  |  | 0.641 |
| No O2 (i) | 598 | 16.4% | 22 | 8.8% | 9 | 12.2% |  |
| Non-invasive O2 (ii) | 1494 | 40.4% | 86 | 34.5% | 27 | 36.5% |  |
| CPAP (iii) | 802 | 21.8% | 55 | 22.1% | 12 | 16.2% |  |
| IMV (iv) | 613 | 17.2% | 85 | 34.1% | 26 | 35.1% |  |
| CXR |  |  |  |  |  |  | <0.001 |
| Normal | 1151 | 28.8% | 29 | 11.6% | 29 | 39.2% |  |
| Other | 261 | 5.9% | 14 | 5.6% | <5 | - |  |
| Indeterminate | 63 | 2.1% | 59 | 23.7% | <5 | - |  |
| Extensive involvement | 40 | 0.9% | 39 | 15.7% | <5 | - |  |
| Suggestive fibrosis | 48 | 1.5% | 42 | 16.9% | 6 | 8.1% |  |
| Missing | 2139 | 60.7% | 66 | 26.5% | 30 | 40.5% |  |
| CT |  |  |  |  |  |  | <0.001 |
| Performed | 422 | 11.4% | 67 | 26.9% | 74 | 100.0% |  |
| PSQ: cough/breathless |  |  |  |  |  |  | 0.879 |
| Present - worsen | 827 | 33.6% | 101 | 40.6% | 32 | 43.2% |  |
| Present - no change | 306 | 12.3% | 24 | 9.6% | 7 | 9.5% |  |
| Not present/improved | 357 | 14.4% | 32 | 12.9% | 12 | 16.2% |  |
| Missing | 2212 | 39.7% | 92 | 36.9% | 23 | 31.1% |  |
| ppFVC |  |  |  |  |  |  | 0.310 |
| 80%+ | 678 | 28.0% | 86 | 34.5% | 23 | 31.1% |  |
| <80% | 260 | 10.7% | 52 | 20.9% | 9 | 12.2% |  |
| Missing | 2764 | 61.3% | 111 | 44.6% | 42 | 56.8% |  |
| ppDLco |  |  |  |  |  |  | <0.001 |
| 80%+ | 246 | 10.2% | 11 | 4.4% | 16 | 21.6% |  |
| <80% | 135 | 5.6% | 126 | 50.6% | 9 | 12.2% |  |
| Missing | 3321 | 84.3% | 112 | 45.0% | 49 | 66.2% |  |

Small numbers <5 have been suppressed. Chi-squared (χ²) performed on non-missing categories. IMD: index of multiple deprivation in quintiles, BMI: body mass index, WHO: modified World Health Organisation severity score, CXR: chest X-ray, CT: computed tomography – chest, PSQ: Patient Symptom Questionnaire, ppFVC: percent predicted forced vital capacity, ppDLco: percent predicted diffusion capacity across the lung for carbon monoxide.

**Supplementary Table 3: Prevalence estimates and sensitivity of priors**

| Model | Posterior mean (%) | 95% CrI | Prior | a | b | DIC |
| --- | --- | --- | --- | --- | --- | --- |
| 1 | 7.42 | 6.55; 8.34 | Uniform | 1 | 1 | 9.33 |
| 1-i | 7.39 | 6.50; 8.29 | Jeffreys | 0.5 | 0.5 | 9.28 |
| 1-ii | 5.71 | 5.03; 6.41 | Sceptical | 1 | 1000 | 25.75 |
| 1-iii | 6.47 | 5.70; 7.25 | Power | 1 | 1000 | 13.94 |
| 2 | 9.93 | 8.69; 11.25 | Uniform | 1 | 1 | 9.13 |
| 2-i | 9.88 | 8.63; 11.20 | Jeffreys | 0.5 | 0.5 | 9.20 |
| 2-ii | 6.73 | 5.85; 7.67 | Sceptical | 1 | 1000 | 38.98 |
| 2-iii | 8.03 | 7.01; 9.11 | Power | 1 | 1000 | 18.46 |

Posterior mean and 95% credibility interval for prevalence of suspected Post COVID ILDam in hospitalised participants (Model 1; Tier 2 sensitivity 2) using uniform priors and in sensitivity with Jeffreys non-informative (i), sceptical informative priors (ii) and sceptical informative priors with power weighting (iii). Beta prior distributions defined using cases (a) and non-cases (b). Deviance information criterion (DIC) presented to interpret model.

**Supplementary Figure 1. Histograms of follow-up time.**

Where CTs were performed and linked. The time from discharge to follow-up visit in the interim cohort (A) and in those with CT scored (B) are plotted. Time between CT date and discharge date (C), and CT date to follow-up visit (D) are plotted.

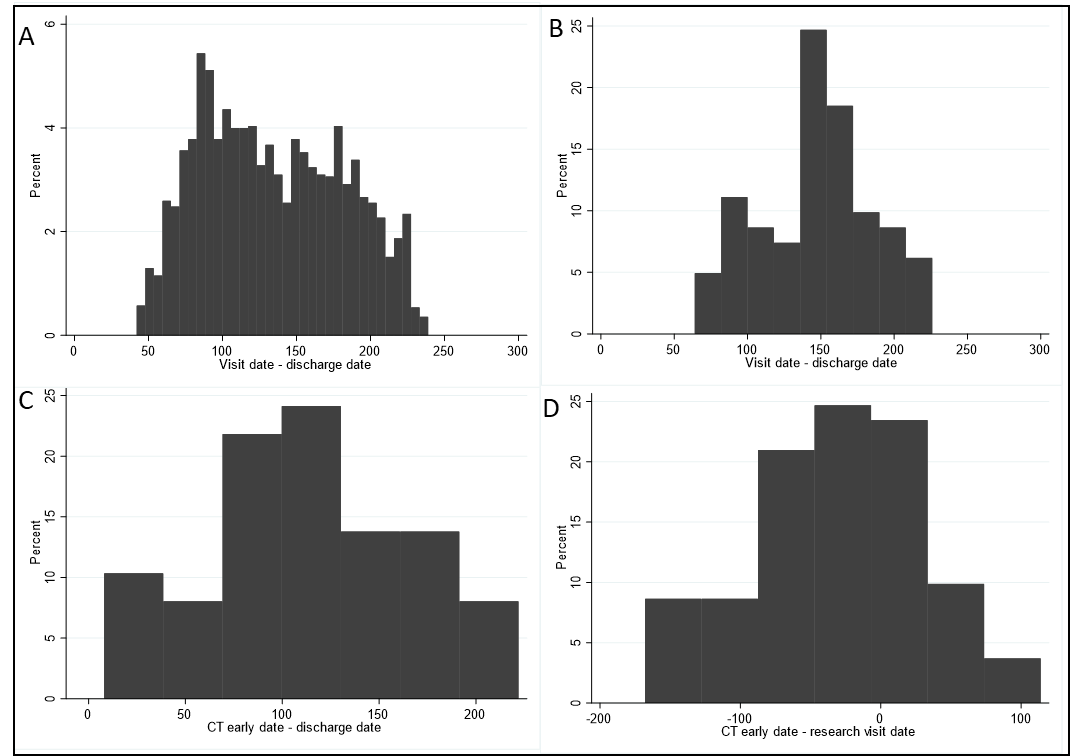

**Supplementary Figure 2. Convergence traces**

Bayes convergence diagnostics provided for prevalence of suspected Post-COVID ILDam in hospitalised participants <240 days with non-informative flat priors (Model 1), and sensitivity based on cases in Tier 2 alone (Model 2).

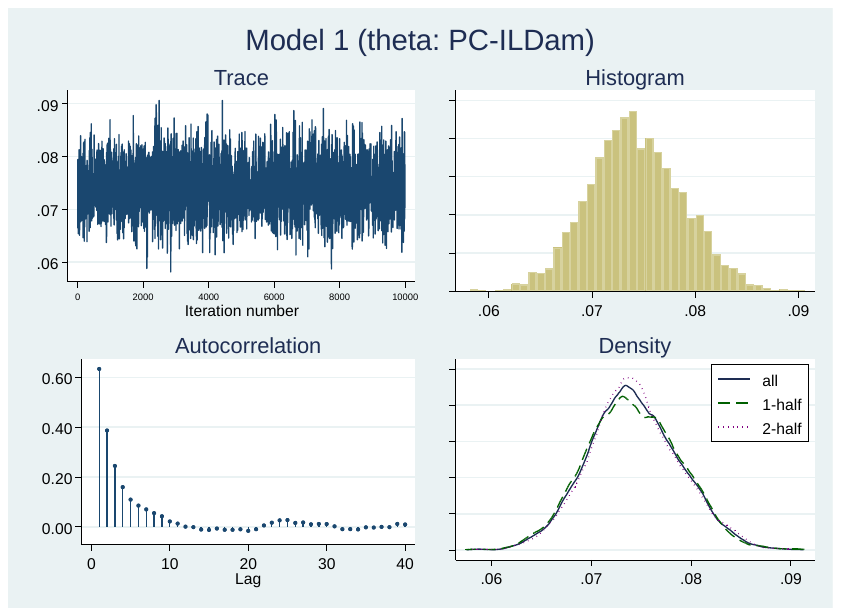

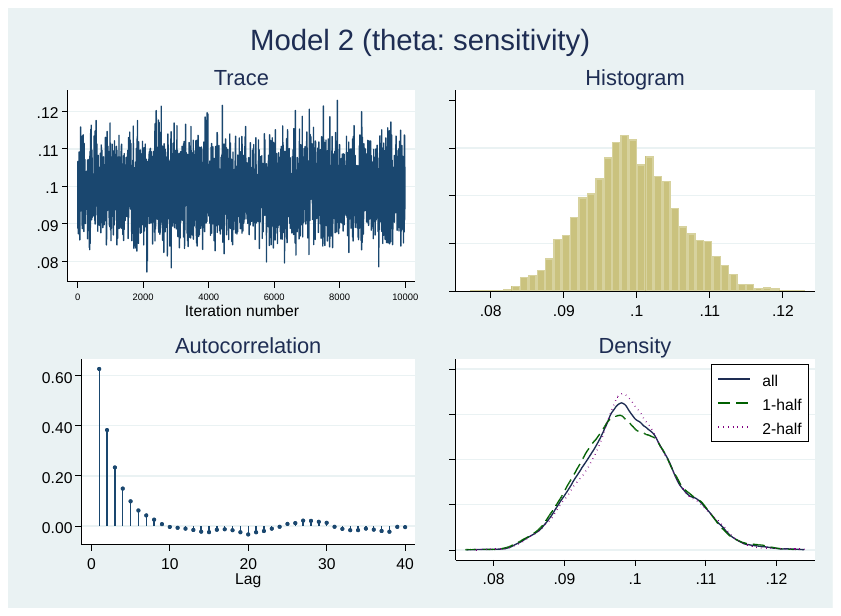

Supplementary Document PHOSP members and affiliations

| **Surname** | **Initial** | **Primary Affiliated Organisation** |
| --- | --- | --- |
| Abel | K | University of Manchester |
| Adamali | H | North Bristol NHS Trust & University of Bristol |
| Adeloye | D | University of Edinburgh |
| Adeyemi | O | King’s College Hospital NHS Foundation Trust |
| Adrego | R | King’s College Hospital NHS Foundation Trust |
| Aguilar Jimenez | L A | Guy’s and St Thomas’ NHS Foundation Trust |
| Ahmad | S | Royal Free London NHS Foundation Trust |
| Ahmad Haider | N | University Hospital Birmingham NHS Foundation Trust |
| Ahmed | R | Stroke Association |
| Ahwireng | N | University College London Hospital |
| Ainsworth | M | Oxford University Hospitals NHS Foundation Trust |
| Al-Sheklly | B | Manchester University NHS Foundation Trust |
| Alamoudi | A | Oxford University Hospitals NHS Foundation Trust |
| Ali | M | St George’s University Hospitals NHS Foundation Trust |
| Aljaroof | M | University Hospitals of Leicester NHS Trust |
| All | AM | Liverpool University Hospitals NHS Foundation Trust |
| Allan | L | University of Exeter |
| Allen | R J | University of Leicester |
| Allerton | L | Liverpool University Hospitals NHS Foundation Trust |
| Allsop | L | Sherwood Forest Hospitals NHS Foundation Trust |
| Almeida | P | Nottingham University Hospitals NHS Trust |
| Altmann | D | Imperial College London |
| Alvarez Corral | M | Hampshire Hospitals NHS Foundation Trust |
| Anderson | D | NHS Greater Glasgow and Clyde Health Board |
| Antoniades | C | University of Oxford |
| Arbane | G | Guy’s and St Thomas’ NHS Foundation Trust |
| Arias | A | Hampshire Hospitals NHS Foundation Trust |
| Armour | C | Belfast Health & Social Care Trust |
| Armstrong | L | Airedale NHS Foundation Trust |
| Armstrong | N | University Hospitals of Leicester NHS Trust |
| Arnold | D | North Bristol NHS Trust |
| Arnold | H | University Hospitals of Leicester NHS Trust |
| Ashish | A | Wrightington Wigan and Leigh NHS trust |
| Ashworth | A | Leeds Teaching Hospitals |
| Ashworth | M | University of Liverpool |
| Aslani | S | University College London |
| Assefa-Kebede | H | King’s College Hospital NHS Foundation Trust |
| Atkin | C | University Hospital Birmingham NHS Foundation Trust |
| Atkin | P | Hull University Teaching Hospitals NHS Trust |
| Aul | R | St George's University Hospitals NHS Foundation Trust |
| Aung | H | University Hospitals of Leicester NHS Trust |
| Austin | L | East Kent Hospitals University NHS Foundation Trust |
| Avram | C | Manchester University NHS Foundation Trust |
| Ayoub | A | Newcastle upon Tyne Hospitals NHS Foundation Trust |
| Babores | M | East Cheshire NHS Trust |
| Baggott | R | University Hospital Birmingham NHS Foundation Trust |
| Bagshaw | J | Sheffield Teaching NHS Foundation Trust |
| Baguley | D | University of Nottingham |
| Bailey | L | Wirral University Teaching Hospital |
| Baillie | J K | Roslin Institute, University of Edinburgh, Edinburgh |
| Bain | S | University of Swansea |
| Bakali | M | University Hospitals of Leicester NHS Trust |
| Bakau | M | University Hospitals of Leicester NHS Trust |
| Baldry | E | University Hospitals of Leicester NHS Trust |
| Baldwin | D | University of Southampton |
| Ballard | C | University of Exeter |
| Bang | B | University College London Hospital |
| Barker | R E | Royal Brompton and Harefield Clinical Group, Guy’s and St Thomas’ NHS Foundation trust. |
| Barman | L | York & Scarborough NHS Foundation Trust |
| Barratt | S | North Bristol NHS Trust |
| Barrett | F | NHS Highland |
| Basire | D | University College London Hospital |
| Basu | N | NHS Greater Glasgow and Clyde Health Board |
| Bates | M | University Hospital Birmingham NHS Foundation Trust |
| Bates | A | University of Oxford |
| Batterham | R | University College London |
| Baxendale | H | Royal Papworth Hospital NHS Foundation Trust |
| Bayes | H | NHS Greater Glasgow and Clyde Health Board |
| Beadsworth | M | Liverpool University Hospitals NHS Foundation Trust |
| Beckett | P | University Hospitals of Derby and Burton |
| Beggs | M | University of Oxford |
| Begum | M | Sheffield Teaching NHS Foundation Trust |
| Beirne | P | Leeds Teaching Hospitals |
| Bell | D | NHS Lanarkshire |
| Bell | R | University College London |
| Bennett | K | Sherwood Forest Hospitals NHS Foundation Trust |
| Beranova | E | East Kent Hospitals University NHS Foundation Trust |
| Bermperi | A | Cambridge University Hospitals NHS Foundation Trust |
| Berridge | A | Liverpool University Hospitals NHS Foundation Trust |
| Berry | C | NHS Greater Glasgow and Clyde Health Board |
| Betts | S | Guy’s and St Thomas’ NHS Foundation Trust |
| Bevan | E | Hampshire Hospitals NHS Foundation Trust |
| Bhui | K | University of Oxford |
| Bingham | M | University of Leicester |
| Birchall | K | Sheffield Teaching NHS Foundation Trust |
| Bishop | L | Loughborough University |
| Bisnauthsing | K | Guy’s and St Thomas’ NHS Foundation Trust |
| Blaikely | J | Manchester University NHS Foundation Trust |
| Bloss | A | Oxford University Hospitals NHS Foundation Trust |
| Bolger | A | Betsi Cadwallader University Health Board |
| Bolton | C E | Nottingham University Hospitals NHS Trust |
| Bonnington | J | Nottingham University Hospitals NHS Trust |
| Botkai | A | University Hospital Birmingham NHS Foundation Trust |
| Bourne | C | University Hospitals of Leicester NHS Trust |
| Bourne | M | University Hospitals of Leicester NHS Trust |
| Bramham | K | King's College London |
| Brear | L | Bradford Teaching Hospitals NHS Foundation Trust |
| Breen | G | South London and Maudsley NHS Foundation Trust |
| Breeze | J | King’s College Hospital NHS Foundation Trust |
| Briggs | A | London School of Hygiene & Tropical Medicine |
| Bright | E | Whittington Health NHS Trust |
| Brightling | C E | University Hospitals of Leicester NHS Trust |
| Brill | S | Royal Free London NHS Foundation Trust |
| Brindle | K | Hull University Teaching Hospitals NHS Trust |
| Broad | L | Cardiff and Vale University Healthy Board |
| Broadley | A | Yeovil District Hospital NHS Foundation Trust |
| Brookes | C | York & Scarborough NHS Foundation Trust |
| Broome | M | University of Birmingham |
| Brown | A | NHS Greater Glasgow and Clyde Health Board |
| Brown | J | Liverpool University Hospitals NHS Foundation Trust |
| Brown | J S | University College London Hospital |
| Brown | M | King's College London |
| Brown | V | Belfast Health & Social Care Trust |
| Brugha | T | University of Leicester |
| Brunskill | N | University Hospitals of Leicester NHS Trust |
| Buch | M | Manchester University NHS Foundation Trust |
| Buckley | P | Sherwood Forest Hospitals NHS Foundation Trust |
| Bularga | A | BHF Centre for Cardiovascular Science |
| Bullmore | E | University of Cambridge |
| Burden | L | Imperial College Healthcare NHS Trust |
| Burdett | T | Harrogate and District NHD Foundation Trust |
| Burn | D | Newcastle University |
| Burns | G | Newcastle upon Tyne Hospitals NHS Foundation Trust |
| Burns | A | Oxford University Hospitals NHS Foundation Trust |
| Busby | J | Queen's University Belfast |
| Butcher | R | Sheffield Teaching NHS Foundation Trust |
| Butt | A | Tameside and Glossop Integrated Care NHS Foundation Trust |
| Byrne | S | King’s College Hospital NHS Foundation Trust |
| Cairns | P | University Hospitals of Leicester NHS Trust |
| Calder | P C | University of Southampton |
| Calvelo | E | Imperial College Healthcare NHS Trust |
| Carborn | H | Sheffield Teaching NHS Foundation Trust |
| Card | B | Imperial College Healthcare NHS Trust |
| Carr | C | Imperial College Healthcare NHS Trust |
| Carr | L | University Hospitals of Leicester NHS Trust |
| Carson | G | University of Oxford, Nuffield Department of Medicine |
| Carter | P | Oxford University Hospitals NHS Foundation Trust |
| Casey | A | University Hospital Birmingham NHS Foundation Trust |
| Cassar | M | Oxford University Hospitals NHS Foundation Trust |
| Cavanagh | J | University of Glasgow |
| Chablani | M | United Lincolnshire Hospitals NHS Trust |
| Chalder | T | Department of Psychological Medicine, King's College London |
| Chalmers | J D | NHS Tayside & University of Dundee |
| Chambers | R C | University College London Hospital |
| Chan | F | Sheffield Teaching NHS Foundation Trust |
| Channon | K M | University of Oxford |
| Chapman | K | Sheffield Teaching NHS Foundation Trust |
| Charalambou | A | University Hospitals of Leicester NHS Trust |
| Chaudhuri | N | University Hospital of South Manchester NHS Foundation Trust |
| Checkley | A | University College London Hospital |
| Chen | J | Oxford University Hospitals NHS Foundation Trust |
| Cheng | Y | Sheffield Teaching NHS Foundation Trust |
| Chetham | L | Sheffield Teaching NHS Foundation Trust |
| Childs | C | University Hospital Southampton NHS Foundation Trust |
| Chilvers | E R | Imperial College Healthcare NHS Trust |
| Chinoy | H | University of Manchester |
| Chiribiri | A | Kings College Hospital, Guys and St Thomas NHS FT |
| Chong-James | K | Barts Health NHS Trust |
| Choudhury | G | NHS Lothian & University of Edinburgh |
| Choudhury | N | Manchester University NHS Foundation Trust |
| Chowienczyk | P | School of Cardiovascular Medicine & Sciences. King’s College London |
| Christie | C | University Hospitals of Leicester NHS Trust |
| Chrystal | M | Nottingham University Hospitals NHS Trust |
| Clark | D | University of Oxford |
| Clark | C | Sheffield Teaching NHS Foundation Trust |
| Clarke | J | Leeds Teaching Hospitals |
| Clohisey | S | NHS Lothian |
| Coakley | G | Lewisham & Greenwich NHS Trust |
| Coburn | Z | Sheffield Teaching NHS Foundation Trust |
| Coetzee | S | Hywel Dda University Health Board |
| Cole | J | Sheffield Teaching NHS Foundation Trust |
| Coleman | C | University of Nottingham |
| Conneh | F | Oxford University Hospitals NHS Foundation Trust |
| Connell | D | NHS Tayside |
| Connolly | B | Queen's University Belfast |
| Connor | L | Swansea Bay University Health Board |
| Cook | A | Swansea Bay University Health Board |
| Cooper | B | University Hospital Birmingham NHS Foundation Trust |
| Cooper | J | Wrightington Wigan and Leigh NHS trust |
| Cooper | S | Liverpool University Hospitals NHS Foundation Trust |
| Copeland | D | Imperial College Healthcare NHS Trust |
| Cosier | T | East Kent Hospitals University NHS Foundation Trust |
| Coulding | M | Tameside and Glossop Integrated Care NHS Foundation Trust |
| Coupland | C | Leeds Teaching Hospitals |
| Cox | E | University of Nottingham |
| Craig | T | Belfast Health & Social Care Trust |
| Crisp | P | Whittington Health NHS Trust |
| Cristiano | D | Royal Brompton and Harefield Clinical Group, Guy’s and St Thomas’ NHS Foundation trust. |
| Crooks | M G | Hull University Teaching Hospitals NHS Trust |
| Cross | A | Liverpool University Hospitals NHS Foundation Trust |
| Cruz | I | Cambridge University Hospitals NHS Foundation Trust |
| Cullinan | P | Imperial College Healthcare NHS Trust |
| Cuthbertson | D | University of Liverpool |
| Daines | L | Usher Institute, University of Edinburgh, Edinburgh, United Kingdom |
| Dalton | M | Leeds Teaching Hospitals |
| Daly | P | Imperial College Healthcare NHS Trust |
| Daniels | A | The Rotherham NHS Foundation Trust |
| Dark | P | Salford Royal NHS Foundation Trust |
| Dasgin | J | University Hospital Birmingham NHS Foundation Trust |
| David | A | University College London |
| David | C | Barts Health NHS Trust |
| Davies | E | Cwm Taf Morgannwg University Health Board |
| Davies | F | Betsi Cadwallader University Health Board |
| Davies | G | Newcastle upon Tyne Hospitals NHS Foundation Trust |
| Davies | G A | Swansea Bay University Health Board |
| Davies | K | Hywel Dda University Health Board |
| Davies | M | University Hospitals of Leicester NHS Trust |
| Dawson | J | Borders General Hospital, NHS Borders |
| Daynes | E | University Hospitals of Leicester NHS Trust |
| De Soyza | A | Newcastle upon Tyne Hospitals NHS Foundation Trust |
| Deakin | B | University of Manchester |
| Deans | A | NHS Lothian & University of Edinburgh |
| Deas | C | NHS Tayside & University of Dundee |
| Deery | J | East Kent Hospitals University NHS Foundation Trust |
| Defres | S | Liverpool University Hospitals NHS Foundation Trust |
| Dell | A | Aneurin Bevan University Health Board |
| Dempsey | K | Cambridge University Hospitals NHS Foundation Trust |
| Denneny | E | University College London |
| Dennis | J | University of Exeter Medical School |
| Dewar | A | Guy’s and St Thomas’ NHS Foundation Trust |
| Dharmagunawardena | R | Whittington Health NHS Trust |
| Diar-Bakerly | N | Salford Royal NHS Foundation Trust |
| Dickens | C | University Hospitals of Derby and Burton |
| Dipper | A | North Bristol NHS Trust & University of Bristol |
| Diver | S | University Hospitals of Leicester NHS Trust |
| Diwanji | S N | London North West University Healthcare NHS Trust |
| Dixon | M | Sheffield Teaching NHS Foundation Trust |
| Djukanovic | R | University Hospital Southampton NHS Foundation Trust |
| Dobson | H | Alzheimer's Research UK |
| Dobson | S L | Liverpool University Hospitals NHS Foundation Trust |
| Docherty | A B | NHS Lothian |
| Donaldson | A | NHS Highland |
| Dong | T | Oxford University Hospitals NHS Foundation Trust |
| Dormand | N | Royal Brompton and Harefield Clinical Group, Guy’s and St Thomas’ NHS Foundation trust. |
| Dougherty | A | NHS Greater Glasgow and Clyde Health Board |
| Dowling | R | University Hospitals of Leicester NHS Trust |
| Drain | S | Belfast Health & Social Care Trust |
| Draxlbauer | K | University Hospital Birmingham NHS Foundation Trust |
| Drury | K | Hull University Teaching Hospitals NHS Trust |
| Dulawan | P | King’s College Hospital NHS Foundation Trust |
| Dunleavy | A | St George’s University Hospitals NHS Foundation Trust |
| Dunn | S | North Bristol NHS Trust & University of Bristol |
| Earley | J | Liverpool University Hospitals NHS Foundation Trust |
| Easom | N | Hull University Teaching Hospitals NHS Trust |
| Echevarria | C | Newcastle upon Tyne Hospitals NHS Foundation Trust |
| Edwards | S | University Hospitals of Leicester NHS Trust |
| Edwardson | C | University Hospitals of Leicester NHS Trust |
| El-Taweel | H | Borders General Hospital, NHS Borders |
| Elliott | A | NHS Tayside & University of Dundee |
| Elliott | K | York & Scarborough NHS Foundation Trust |
| Ellis | Y | Health & Care Research Wales |
| Elmer | A | Cambridge University Hospitals NHS Foundation Trust |
| Elneima | O | University Hospitals of Leicester NHS Trust |
| Evans | D | Salford Royal NHS Foundation Trust |
| Evans | H | University Hospitals of Leicester NHS Trust |
| Evans | J | University of Bristol |
| Evans | R | University College London Hospital |
| Evans | R A | University Hospitals of Leicester NHS Trust |
| Evans | R I | Oxford University Hospitals NHS Foundation Trust |
| Evans | T | Cardiff and Vale University Healthy Board |
| Evenden | C | Cwm Taf Morgannwg University Health Board |
| Evison | L | Imperial College Healthcare NHS Trust |
| Fabbri | L | University of Nottingham |
| Fairbairn | S | Aneurin Bevan University Health Board |
| Fairman | A | Sheffield Teaching NHS Foundation Trust |
| Fallon | K | NHS Greater Glasgow and Clyde Health Board |
| Faluyi | D | Manchester University NHS Foundation Trust |
| Favager | C | Leeds Teaching Hospitals |
| Fayzan | T | Imperial College Healthcare NHS Trust |
| Featherstone | J | Harrogate and District NHD Foundation Trust |
| Felton | T | Manchester University NHS Foundation Trust |
| Finch | J | University Hospitals of Leicester NHS Trust |
| Finney | S | University of Leicester |
| Finnigan | J | Sheffield Teaching NHS Foundation Trust |
| Finnigan | L | University of Sheffield |
| Fisher | H | Newcastle upon Tyne Hospitals NHS Foundation Trust |
| Fletcher | S | University Hospital Southampton NHS Foundation Trust |
| Flockton | R | Hull University Teaching Hospitals NHS Trust |
| Flynn | M | Sherwood Forest Hospitals NHS Foundation Trust |
| Foot | H | Sheffield Teaching NHS Foundation Trust |
| Foote | D | Sheffield Teaching NHS Foundation Trust |
| Ford | A | Sheffield Teaching NHS Foundation Trust |
| Forton | D | St George's University Hospitals NHS Foundation Trust |
| Fraile | E | The Great Western Hospital Foundation Trust |
| Francis | C | Newcastle upon Tyne Hospitals NHS Foundation Trust |
| Francis | R | Stroke Association |
| Francis | S | Nottingham University |
| Frankel | A | Imperial College London |
| Fraser | E | Oxford University Hospitals NHS Foundation Trust |
| Free | R | University of Leicester |
| French | N | Liverpool University Hospitals NHS Foundation Trust |
| Fu | X | University of Oxford |
| Fuld | J | Cambridge University Hospitals NHS Foundation Trust |
| Furniss | J | NHS Lothian & University of Edinburgh |
| Garner | L | Royal Papworth Hospital NHS Foundation Trust |
| Gautam | N | University Hospital Birmingham NHS Foundation Trust |
| Geddes | J R | Oxford University Hospitals NHS Foundation Trust |
| George | J | NHS Tayside & University of Dundee |
| George | P | Royal Brompton and Harefield Clinical Group, Guy’s and St Thomas’ NHS Foundation trust. |
| Gibbons | M | Royal Devon and Exeter NHS Trust |
| Gill | M | Sherwood Forest Hospitals NHS Foundation Trust |
| Gilmour | L | NHS Greater Glasgow and Clyde Health Board |
| Gleeson | F | Oxford University Hospitals NHS Foundation Trust |
| Glossop | J | Leeds Teaching Hospitals |
| Glover | S | University Hospitals of Leicester NHS Trust |
| Goodman | N | University Hospitals of Leicester NHS Trust |
| Goodwin | C | Sherwood Forest Hospitals NHS Foundation Trust |
| Gooptu | B | University Hospitals of Leicester NHS Trust |
| Gordon | H | Imperial College Healthcare NHS Trust |
| Gorsuch | T | Manchester University NHS Foundation Trust |
| Greatorex | M | Sherwood Forest Hospitals NHS Foundation Trust |
| Greenhaff | P L | Nottingham University Hospitals NHS Trust |
| Greenhalf | W | Liverpool University Hospitals NHS Foundation Trust |
| Greenhalgh | A | Newcastle upon Tyne Hospitals NHS Foundation Trust |
| Greening | N J | University Hospitals of Leicester NHS Trust |
| Greenwood | J | Leeds Teaching Hospitals |
| Gregory | H | Sherwood Forest Hospitals NHS Foundation Trust |
| Gregory | R | Sheffield Teaching NHS Foundation Trust |
| Grieve | D | NHS Greater Glasgow and Clyde Health Board |
| Griffin | D | Hampshire Hospitals NHS Foundation Trust |
| Griffiths | L | York & Scarborough NHS Foundation Trust |
| Guerdette | A-M | Kettering General Hospital NHS Trust |
| Guillen Guio | B | University of Leicester |
| Gummadi | M | Royal Brompton and Harefield Clinical Group, Guy’s and St Thomas’ NHS Foundation trust. |
| Gupta | A | Nottingham University Hospitals NHS Trust |
| Gurram | S | London North West University Healthcare NHS Trust |
| Guthrie | E | University of Leeds |
| Guy | [Z](mailto:) | York & Scarborough NHS Foundation Trust |
| H Henson | H | Airedale NHS Foundation Trust |
| Hadley | K | University Hospitals of Leicester NHS Trust |
| Haggar | A | Betsi Cadwallader University Health Board |
| Hainey | K | Liverpool University Hospitals NHS Foundation Trust |
| Hairsine | B | Airedale NHS Foundation Trust |
| Haldar | P | University Hospitals of Leicester NHS Trust |
| Hall | I | University of Nottingham |
| Hall | L | Leeds Teaching Hospitals |
| Halling-Brown | M | Royal Surrey NHS Foundation Trust |
| Hamil | R | NHS Lanarkshire |
| Hancock | A | Cwm Taf Morgannwg University Health Board |
| Hancock | K | Cwm Taf Morgannwg University Health Board |
| Hanley | N A | Manchester University NHS Foundation Trust |
| Haq | S | Imperial College Healthcare NHS Trust |
| Hardwick | H E | Liverpool University Hospitals NHS Foundation Trust |
| Hardy | E | Salford Royal NHS Foundation Trust |
| Hardy | T | Leeds Teaching Hospitals |
| Hargadon | B | University Hospitals of Leicester NHS Trust |
| Harrington | K | Sheffield Teaching NHS Foundation Trust |
| Harris | E | Chesterfield Royal Hospital NHS Trust |
| Harris | V C | University Hospitals of Leicester NHS Trust |
| Harrison | E M | NHS Lothian |
| Harrison | P | Oxford University Hospitals NHS Foundation Trust |
| Hart | N | Guy’s and St Thomas’ NHS Foundation Trust |
| Harvey | A | Salford Royal NHS Foundation Trust |
| Harvey | M | University Hospital Southampton NHS Foundation Trust |
| Harvie | M | University of Manchester |
| Haslam | L | Sheffield Teaching NHS Foundation Trust |
| Havinden-Williams | M | Oxford University Hospitals NHS Foundation Trust |
| Hawkes | J | Liverpool University Hospitals NHS Foundation Trust |
| Hawkings | N | Aneurin Bevan University Health Board |
| Haworth | J | Aneurin Bevan University Health Board |
| Hayday | A | Kings College Hospital NHS Foundation Trust |
| Haynes | M | Cardiff and Vale University Healthy Board |
| Hazeldine | J | University Hospital Birmingham NHS Foundation Trust |
| Hazelton | T | East Kent Hospitals University NHS Foundation Trust |
| Heaney | L G | Belfast Health & Social Care Trust |
| Heeley | C | Sherwood Forest Hospitals NHS Foundation Trust |
| Heeney | J L | University of Cambridge |
| Heightman | M | University College London Hospital |
| Henderson | M | University of Leeds |
| Hesselden | L | Sheffield Teaching NHS Foundation Trust |
| Hewitt | M | Kettering General Hospital NHS Trust |
| Highett | V | Liverpool University Hospitals NHS Foundation Trust |
| Hillman | T | University College London Hospital |
| Hingorani | A | University College London |
| Hiwot | T | University Hospital Birmingham NHS Foundation Trust |
| Ho | L P | Oxford University Hospitals NHS Foundation Trust |
| Hoare | A | King’s College Hospital NHS Foundation Trust |
| Hoare | M | Aneurin Bevan University Health Board |
| Hockridge | J | Sheffield Teaching NHS Foundation Trust |
| Hogarth | P | Newcastle upon Tyne Hospitals NHS Foundation Trust |
| Holbourn | A | Sheffield Teaching NHS Foundation Trust |
| Holden | S | University Hospital Birmingham NHS Foundation Trust |
| Holdsworth | L | Hull University Teaching Hospitals NHS Trust |
| Holgate | D | Salford Royal NHS Foundation Trust |
| Holland | M | East Cheshire NHS Trust |
| Holloway | L | Sherwood Forest Hospitals NHS Foundation Trust |
| Holmes | K | NIHR Office for Clinical Research Infrastructure |
| Holmes | M | Sherwood Forest Hospitals NHS Foundation Trust |
| Holroyd-Hind | B | Sheffield Teaching NHS Foundation Trust |
| Holt | L | Sheffield Teaching NHS Foundation Trust |
| Hormis | A | The Rotherham NHS Foundation Trust |
| Horsley | A | Manchester University NHS Foundation Trust |
| Hosseini | A | Nottingham University Hospitals NHS Trust |
| Hotopf | M | South London and Maudsley NHS Foundation Trust |
| Houchen | L | University Hospitals of Leicester NHS Trust |
| Howard | K | York & Scarborough NHS Foundation Trust |
| Howard | L | Imperial College London |
| Howell | A | Sheffield Teaching NHS Foundation Trust |
| Hufton | E | University of Nottingham |
| Hughes | A D | University College London |
| Hughes | J | Newcastle upon Tyne Hospitals NHS Foundation Trust |
| Hughes | R | Hywel Dda University Health Board |
| Humphries | A | Leeds Teaching Hospitals & University of Leeds |
| Huneke | N | University of Southampton |
| Hurditch | E | Sheffield Teaching NHS Foundation Trust |
| Hurst | J | Royal Free London NHS Foundation Trust |
| Husain | M | University of Oxford |
| Hussell | T | Manchester University NHS Foundation Trust |
| Hutchinson | J | Sherwood Forest Hospitals NHS Foundation Trust |
| Ibrahim | W | University Hospitals of Leicester NHS Trust |
| Ilyas | F | Sheffield Teaching NHS Foundation Trust |
| Ingham | J | The Rotherham NHS Foundation Trust |
| Ingram | L | University Hospitals of Leicester NHS Trust |
| Ionita | D | York & Scarborough NHS Foundation Trust |
| Isaacs | K | University Hospital Birmingham NHS Foundation Trust |
| Ismail | K | King's College London |
| Jackson | T | University Hospital Birmingham NHS Foundation Trust |
| Jacob | J | University College London Hospital |
| James | W Y | Barts Health NHS Trust |
| Jarman | C | Sheffield Teaching NHS Foundation Trust |
| Jarrold | I | Asthma UK BLF |
| Jarvis | H | Royal Free London NHS Foundation Trust |
| Jastrub | R | University College London Hospital |
| Jayaraman | B | North Middlesex University Hospital NHS Trust |
| Jenkins | R G | Imperial College London |
| Jezzard | P | Oxford University Hospitals NHS Foundation Trust |
| Jiwa | K | Newcastle upon Tyne Hospitals NHS Foundation Trust |
| Johnson | C | Royal Papworth Hospital NHS Foundation Trust |
| Johnson | S | University of Nottingham |
| Johnston | D | Imperial College London |
| Jolley | C J | King’s College Hospital NHS Foundation Trust |
| Jones | D | University of Leicester |
| Jones | G | Newcastle upon Tyne Hospitals NHS Foundation Trust |
| Jones | H | Cambridge University Hospitals NHS Foundation Trust |
| Jones | I | Cardiff Univeristy, National Centre for Mental Health |
| Jones | L | Cardiff and Vale University Healthy Board |
| Jones | M | University Hospital Southampton NHS Foundation Trust |
| Jones | S | Action for Pulmonary Fibrosis |
| Jose | S | Cambridge University Hospitals NHS Foundation Trust |
| Kabir | T | McPin Foundation |
| Kaltsakas | G | Guy’s and St Thomas’ NHS Foundation Trust |
| Kamwa | V | University Hospital Birmingham NHS Foundation Trust |
| Kanellakis | N | Oxford University Hospitals NHS Foundation Trust |
| Kaprowska | s | Liverpool University Hospitals NHS Foundation Trust |
| Kausar | Z | Manchester University NHS Foundation Trust |
| Keenan | N | East Cheshire NHS Trust |
| Kelly | S | NHS Lothian |
| Kemp | G | University of Liverpool |
| Kerr | S | Roslin Institute, The University of Edinburgh |
| Kerslake | H | Guy’s and St Thomas’ NHS Foundation Trust |
| Key | A L | Liverpool University Hospitals NHS Foundation Trust |
| Khan | F | University of Nottingham |
| Khunti | K | University Hospitals of Leicester NHS Trust |
| Kilroy | S | Tameside and Glossop Integrated Care NHS Foundation Trust |
| King | B | Belfast Health & Social Care Trust |
| King | C | Imperial College Healthcare NHS Trust |
| Kirk | J | Sherwood Forest Hospitals NHS Foundation Trust |
| Kitterick | P | University of Nottingham |
| Klenerman | P | University of Oxford |
| Knibbs | L | Cardiff and Vale University Healthy Board |
| Knight | S | Salford Royal NHS Foundation Trust |
| Knighton | A | King’s College Hospital NHS Foundation Trust |
| Kon | O | Imperial College Healthcare NHS Trust |
| Kon | S | Royal Brompton and Harefield Clinical Group, Guy’s and St Thomas’ NHS Foundation trust. |
| Kon | S S | The Hillingdon Hospitals NHS Foundation Trust |
| Koprowska | S | Liverpool University Hospitals NHS Foundation Trust |
| Korszun | A | Queen Mary University of London |
| Kotanidis | C | University of Oxford, Division of Cardiovascular Medicine |
| Koychev | I | Oxford University Hospitals NHS Foundation Trust |
| Kurasz | C | Airedale NHS Foundation Trust |
| Kurupati | P | Oxford University Hospitals NHS Foundation Trust |
| Laing | C | Royal Free London NHS Foundation Trust |
| Lamlum | H | University of Oxford |
| Landers | G | The Hillingdon Hospitals NHS Foundation Trust |
| Langenberg | C | University of Cambridge |
| Lasserson | D | University of Warwick |
| Lavelle-Langham | L | Liverpool University Hospitals NHS Foundation Trust |
| Lawrie | A | Sheffield Teaching NHS Foundation Trust |
| Lawson | C | University of Leicester |
| Layton | A | Harrogate and District NHD Foundation Trust |
| Lea | A | University Hospitals of Leicester NHS Trust |
| Leavy | O C | University of Leicester |
| Lee | D | University Hospitals of Leicester NHS Trust |
| Lee | J-H | Sheffield Teaching NHS Foundation Trust |
| Lee | E | Sheffield Teaching NHS Foundation Trust |
| Leitch | K | NHS Lanarkshire |
| Lenagh | R | Sheffield Teaching NHS Foundation Trust |
| Lewis | D | University Hospital Birmingham NHS Foundation Trust |
| Lewis | J | Betsi Cadwallader University Health Board |
| Lewis | K | Swansea University |
| Lewis | K E | Hywel Dda University Health Board |
| Lewis | V | Aneurin Bevan University Health Board |
| Lewis-Burke | N | Liverpool University Hospitals NHS Foundation Trust |
| Li | X | University of Oxford |
| Light | T | North Middlesex University Hospital NHS Trust |
| Lightstone | L | Imperial College London |
| Lilaonitkul | W | University College London |
| Lim | L | Royal Free London NHS Foundation Trust |
| Linford | S | Nottingham University Hospitals NHS Trust |
| Lingford-Hughes | A | Imperial College London |
| Lipman | M | University College London Hospital |
| Liyanage | K | Royal Brompton and Harefield Clinical Group, Guy’s and St Thomas’ NHS Foundation trust. |
| Lloyd | A | Betsi Cadwallader University Health Board |
| Logan | S | University College London Hospital |
| Lomas | D | University College London Hospital |
| Lone | N I | Usher Institute, University of Edinburgh |
| Loosley | R | Hywel Dda University Health Board |
| Lord | J M | University Hospital Birmingham NHS Foundation Trust |
| Lota | H | The Hillingdon Hospitals NHS Foundation Trust |
| Lovegrove | W | Sherwood Forest Hospitals NHS Foundation Trust |
| Lucey | A | Aneurin Bevan University Health Board |
| Lukaschuk | E | University of Oxford |
| Lye | A | Sheffield Teaching NHS Foundation Trust |
| Lynch | C | Cwm Taf Morgannwg University Health Board |
| MacDonald | S | University of Glasgow |
| MacGowan | G | Newcastle upon Tyne Hospitals NHS Foundation Trust |
| Macharia | I | Sheffield Teaching NHS Foundation Trust |
| Mackie | J | Royal Papworth Hospital NHS Foundation Trust |
| Macliver | L | NHS Lanarkshire |
| Madathil | S | University Hospital Birmingham NHS Foundation Trust |
| Madzamba | G | Liverpool University Hospitals NHS Foundation Trust |
| Magee | N | Belfast Health & Social Care Trust & Queen's University Belfast |
| Magtoto | M M | Guy’s and St Thomas’ NHS Foundation Trust |
| Mairs | N | Salford Royal NHS Foundation Trust |
| Majeed | N | Salford Royal NHS Foundation Trust |
| Major | E | Belfast Health & Social Care Trust |
| Malein | F | Liverpool University Hospitals NHS Foundation Trust |
| Malim | M | King's College Hospital NHS Foundation Trust |
| Mallison | G | Aneurin Bevan University Health Board |
| Man | W | Imperial College London |
| Mandal | S | Royal Free London NHS Foundation Trust |
| Mangion | K | NHS Greater Glasgow and Clyde Health Board |
| Manisty | C | Barts Heart Centre |
| Manley | R | Betsi Cadwallader University Health Board |
| March | K | Imperial College Healthcare NHS Trust |
| Marciniak | S | Cambridge University Hospitals NHS Foundation Trust |
| Marino | P | Guy’s and St Thomas’ NHS Foundation Trust |
| Mariveles | M | Imperial College Healthcare NHS Trust |
| Marks | M | London School of Hygiene & Tropical Medicine |
| Marouzet | E | University Hospital Southampton NHS Foundation Trust |
| Marsh | S | Liverpool University Hospitals NHS Foundation Trust |
| Marshall | B | University Hospital Southampton NHS Foundation Trust |
| Marshall | M | Sheffield Teaching NHS Foundation Trust |
| Martin | J | Hampshire Hospitals NHS Foundation Trust |
| Martineau | A | Barts Health NHS Trust |
| Martinez | L M | Guy’s and St Thomas’ NHS Foundation Trust |
| Maskell | N | North Bristol NHS Trust & University of Bristol |
| Matila | D | Royal Free London NHS Foundation Trust |
| Matimba-Mupaya | W | Salisbury NHS Foundation Trust |
| Matthews | L | Nottingham University Hospitals NHS Trust |
| Mbuyisa | A | Sheffield Teaching NHS Foundation Trust |
| McAdoo | S | Imperial College London |
| Weir McCall | J | University of Cambridge |
| McAllister-Williams | H | Newcastle University |
| McArdle | A | University of Liverpool |
| McArdle | P | University of Birmingham |
| McAulay | D | Belfast Health & Social Care Trust |
| McCann | G P | University Hospitals of Leicester NHS Trust |
| McCauley | H J C | University Hospitals of Leicester NHS Trust |
| McCormick | J | Tameside and Glossop Integrated Care NHS Foundation Trust |
| McCormick | W | Gateshead NHS Trust |
| McCourt | P | University Hospitals of Leicester NHS Trust |
| McGarvey | L | Belfast Health & Social Care Trust |
| McGee | C | University Hospital Birmingham NHS Foundation Trust |
| Mcgee | K | University Hospital Birmingham NHS Foundation Trust |
| McGinness | J | Belfast Health & Social Care Trust |
| McGlynn | K | University of Oxford |
| McGovern | A | University of Exeter |
| McGuinness | H | Hywel Dda University Health Board |
| McInnes | I B | NHS Greater Glasgow and Clyde Health Board |
| McIntosh | J | Tameside and Glossop Integrated Care NHS Foundation Trust |
| McIvor | E | Betsi Cadwallader University Health Board |
| McIvor | K | Manchester Centre for Clinical Neurosciences, Salford Royal NHS Foundation Trust |
| McLeavey | L | Imperial College Healthcare NHS Trust |
| McMahon | A | Kidney Research UK |
| McMahon | M J | NHS Dumfries and Galloway |
| McMorrow | L | Salford Royal NHS Foundation Trust |
| Mcnally | T | University Hospitals of Leicester NHS Trust |
| McNarry | M | Swansea University |
| McNeill | J | Sheffield Teaching NHS Foundation Trust |
| McQueen | A | Cardiff and Vale University Healthy Board |
| McShane | H | Oxford University Hospitals NHS Foundation Trust |
| Mears | C | Liverpool University Hospitals NHS Foundation Trust |
| Megson | C | Oxford University Hospitals NHS Foundation Trust |
| Megson | S | Sheffield Teaching NHS Foundation Trust |
| Mehta | P | University College London |
| Meiring | J | Sheffield Teaching NHS Foundation Trust |
| Melling | L | Liverpool University Hospitals NHS Foundation Trust |
| Mencias | M | St George’s University Hospitals NHS Foundation Trust |
| Menzies | D | Betsi Cadwallader University Health Board |
| Merida Morillas | M | University College London Hospital |
| Michael | A | Royal Papworth Hospital NHS Foundation Trust |
| Milligan | L | MQ Mental Health Research, |
| Miller | C | University of Manchester |
| Mills | C | Harrogate and District NHD Foundation Trust |
| Mills | N L | BHF Centre for Cardiovascular Science, University of Edinburgh |
| Milner | L | Sheffield Teaching NHS Foundation Trust |
| Misra | S | Sheffield Teaching NHS Foundation Trust |
| Mitchell | J | Imperial College London |
| Mohamed | A | Hywel Dda University Health Board |
| Mohamed | N | Imperial College Healthcare NHS Trust |
| Mohammed | S | NHS Tayside |
| Molyneaux | P L | Imperial College London |
| Monteiro | W | University Hospitals of Leicester NHS Trust |
| Moriera | S | Imperial College Healthcare NHS Trust |
| Morley | A | North Bristol NHS Trust & University of Bristol |
| Morrison | L | North Bristol NHS Trust & University of Bristol |
| Morriss | R | University of Nottingham |
| Morrow | A | NHS Greater Glasgow and Clyde Health Board |
| Moss | A J | University of Leicester |
| Moss | P | University of Birmingham |
| Motohashi | K | University of Oxford |
| Msimanga | N | St George’s University Hospitals NHS Foundation Trust |
| Mukaetova-Ladinska | E | University of Leicester |
| Munawar | U | Imperial College Healthcare NHS Trust |
| Murira | J | Leeds Teaching Hospitals |
| Nanda | U | University Hospitals of Derby and Burton |
| Nassa | H | Aneurin Bevan University Health Board |
| Nasseri | M | The Hillingdon Hospitals NHS Foundation Trust |
| Neal | A | University Hospital Birmingham NHS Foundation Trust |
| Needham | R | Nottingham University Hospitals NHS Trust |
| Neill | P | NHS Dumfries and Galloway |
| Neubauer | S | Oxford University Hospitals NHS Foundation Trust |
| Newby | D E | University of Edinburgh |
| Newell | H | Sheffield Teaching NHS Foundation Trust |
| Newman | T | Sheffield Teaching NHS Foundation Trust |
| Newton-Cox | A | University Hospital Birmingham NHS Foundation Trust |
| Nicholson | T | King's College London |
| Nicoll | D | Oxford University Hospitals NHS Foundation Trust |
| Nolan | C M | Royal Brompton and Harefield Clinical Group, Guy’s and St Thomas’ NHS Foundation trust. |
| Noonan | M J | Liverpool University Hospitals NHS Foundation Trust |
| Norman | C | Sheffield Teaching NHS Foundation Trust |
| Novotny | P | University of Leicester |
| Nunag | J | Imperial College Healthcare NHS Trust |
| Nwafor | L | Sheffield Teaching NHS Foundation Trust |
| Nwanguma | U | Imperial College Healthcare NHS Trust |
| Nyaboko | J | University Hospital Birmingham NHS Foundation Trust |
| O'Donnell | K | University of Glasgow |
| O'Brien | C | Kings College Hospital, Guys and St Thomas NHS FT |
| O’Brien | L | Hywel Dda University Health Board |
| O'Regan | D | Imperial College London |
| Odell | N | Manchester University NHS Foundation Trust |
| Ogg | G | Oxford University Hospitals NHS Foundation Trust |
| Olaosebikan | O | Royal Free London NHS Foundation Trust |
| Oliver | C | Cardiff and Vale University Healthy Board |
| Omar | Z | Hywel Dda University Health Board |
| Openshaw | P J M | National Heart and Lung Institute, Imperial College London, London, United Kingdom |
| Orriss-Dib | L | Imperial College Healthcare NHS Trust |
| Osborne | L | United Lincolnshire Hospitals NHS Trust |
| Osbourne | R | Manchester University NHS Foundation Trust |
| Ostermann | M | Guy’s and St Thomas’ NHS Foundation Trust |
| Overton | C | University of Leicester |
| Owen | J | Hampshire Hospitals NHS Foundation Trust |
| Oxton | J | Salford Royal NHS Foundation Trust |
| Pack | J | Royal Papworth Hospital NHS Foundation Trust |
| Pacpaco | E | Oxford University Hospitals NHS Foundation Trust |
| Paddick | S | Newcastle University |
| Painter | S | Shropshire Community Health NHS Trust |
| Pakzad | A | University College London |
| Palmer | S | Somerset NHS Foundation Trust |
| Papineni | P | London North West University Healthcare NHS Trust |
| Paques | K | Royal Papworth Hospital NHS Foundation Trust |
| Paradowski | K | Cardiff and Vale University Healthy Board |
| Pareek | M | University Hospitals of Leicester NHS Trust |
| Parekh | D | University Hospital Birmingham NHS Foundation Trust |
| Parfrey | H | Royal Papworth Hospital NHS Foundation Trust |
| Pariante | C | King's College London |
| Parker | S | University Hospitals of Leicester NHS Trust |
| Parkes | M | Cambridge University Hospitals NHS Foundation Trust |
| Parmar | J | Royal Papworth Hospital NHS Foundation Trust |
| Patale | S | King’s College Hospital NHS Foundation Trust |
| Patel | B | Royal Brompton and Harefield Clinical Group, Guy’s and St Thomas’ NHS Foundation trust. |
| Patel | M | NHS Lanarkshire |
| Patel | S | Royal Brompton and Harefield Clinical Group, Guy’s and St Thomas’ NHS Foundation trust. |
| Pattenadk | D | Sheffield Teaching NHS Foundation Trust |
| Pavlides | M | Oxford University Hospitals NHS Foundation Trust |
| Payne | S | Hampshire Hospitals NHS Foundation Trust |
| Pearce | L | Gateshead NHS Trust |
| Pearl | J E | University of Leicester |
| Peckham | D | Leeds Teaching Hospitals |
| Pendlebury | J | Salford Royal NHS Foundation Trust |
| Peng | Y | Oxford University Hospitals NHS Foundation Trust |
| Pennington | C | Aneurin Bevan University Health Board |
| Peralta | I | King’s College Hospital NHS Foundation Trust |
| Perkins | E | Hywel Dda University Health Board |
| Peterkin | Z | University Hospital Birmingham NHS Foundation Trust |
| Peto | T | Queen's University Belfast |
| Petousi | N | Oxford University Hospitals NHS Foundation Trust |
| Petrie | J | University of Glasgow |
| Pfeffer | P | Barts Health NHS Trust |
| Phipps | J | Hywel Dda University Health Board |
| Pimm | J | Oxford University Hospitals NHS Foundation Trust |
| Piper Hanley | K | Manchester University NHS Foundation Trust |
| Pius | R | University of Edinburgh |
| Plant | H | University College London Hospital |
| Plein | S | Leeds Teaching Hospitals |
| Plekhanova | T | University of Leicester |
| Plowright | M | Sheffield Teaching NHS Foundation Trust |
| Poinasamy | K | Asthma UK and British Lung Foundation Partnership |
| Polgar | O | Royal Brompton and Harefield Clinical Group, Guy’s and St Thomas’ NHS Foundation trust. |
| Poll | L | Liverpool University Hospitals NHS Foundation Trust |
| Porter | J C | University College London Hospital |
| Porter | J | Sheffield Teaching NHS Foundation Trust |
| Portukhay | S | The Hillingdon Hospitals NHS Foundation Trust |
| Powell | N | King’s College Hospital NHS Foundation Trust |
| Prabhu | A | Hampshire Hospitals NHS Foundation Trust |
| Pratt | J | Liverpool University Hospitals NHS Foundation Trust |
| Price | A | Aneurin Bevan University Health Board |
| Price | C | East Kent Hospitals University NHS Foundation Trust |
| Price | D | Newcastle upon Tyne Hospitals NHS Foundation Trust |
| Price | L | Royal Brompton Hospital |
| Prickett | A | University Hospitals of Leicester NHS Trust |
| Propescu | J | University of Oxford |
| Pugmire | S | Gateshead NHS Trust |
| Quaid | S | London North West University Healthcare NHS Trust |
| Quigley | J | NHS Lanarkshire |
| Quint | J | Imperial College London |
| Qureshi | H | University Hospital Birmingham NHS Foundation Trust |
| Qureshi | I N | University Hospitals of Leicester NHS Trust |
| Radhakrishnan | K | Manchester University NHS Foundation Trust |
| Rahman | N M | Oxford University Hospitals NHS Foundation Trust |
| Ralser | M | Francis Crick Institute |
| Raman | B | Oxford University Hospitals NHS Foundation Trust |
| Ramos | A | King’s College Hospital NHS Foundation Trust |
| Ramos | H | East Kent Hospitals University NHS Foundation Trust |
| Rangeley | J | Leeds Teaching Hospitals |
| Rangelov | B | University College London |
| Ratcliffe | L | University Hospital Birmingham NHS Foundation Trust |
| Ravencroft | P | Sheffield Teaching NHS Foundation Trust |
| Reddington | A | Wirral University Teaching Hospital |
| Reddy | R | Kettering General Hospital NHS Trust |
| Redfearn | H | York & Scarborough NHS Foundation Trust |
| Redwood | D | Somerset NHS Foundation Trust |
| Reed | A | Hampshire Hospitals NHS Foundation Trust |
| Rees | M | Cwm Taf Morgannwg University Health Board |
| Rees | T | Swansea Bay University Health Board |
| Regan | K | Bradford Teaching Hospitals NHS Foundation Trust |
| Reynolds | W | University of Liverpool |
| Ribeiro | C | Cambridge University Hospitals NHS Foundation Trust |
| Richards | A | Hull University Teaching Hospitals NHS Trust |
| Richardson | E | Liverpool University Hospitals NHS Foundation Trust |
| Richardson | M | University of Leicester |
| Rivera-Ortega | P | Manchester University NHD Foundation Trust |
| Roberts | K | Betsi Cadwallader University Health Board |
| Robertson | E | Diabetes UK, University of Glasgow |
| Robinson | E | Wrightington Wigan and Leigh NHS trust |
| Robinson | L | Borders General Hospital, NHS Borders |
| Roche | L | Cwm Taf Morgannwg University Health Board |
| Roddis | C | Sheffield Teaching NHS Foundation Trust |
| Rodger | J | Sheffield Teaching NHS Foundation Trust |
| Ross | A | Imperial College Healthcare NHS Trust |
| Ross | G | Hywel Dda University Health Board |
| Rossdale | J | Guy’s and St Thomas’ NHS Foundation Trust |
| Rostron | A | University of Newcastle |
| Rowe | A | Liverpool University Hospitals NHS Foundation Trust |
| Rowland | A | University Hospitals of Leicester NHS Trust |
| Rowland | J | NHS Tayside & University of Dundee |
| Rowland | M J | Oxford University Hospitals NHS Foundation Trust |
| Rowland-Jones | S L | Sheffield Teaching NHS Foundation Trust |
| Roy | K | University College London Hospital |
| Roy | M | Imperial College Healthcare NHS Trust |
| Rudan | I | University of Edinburgh |
| Russell | R | University Hospitals of Leicester NHS Trust |
| Russell | E | Imperial College Healthcare NHS Trust |
| Saalmink | G | Leeds Teaching Hospitals |
| Sabit | R | Cardiff and Vale University Healthy Board |
| Sage | E K | NHS Highland |
| Samakomva | T | St George’s University Hospitals NHS Foundation Trust |
| Samani | N | University of Leicester |
| Sampson | C | Chesterfield Royal Hospital NHS Trust |
| Samuel | K | Imperial College Healthcare NHS Trust |
| Samuel | R | University Hospital Southampton NHS Foundation Trust |
| Sanderson | A | Barnsley Hospital NHS Foundation Trust |
| Sapey | E | University Hospital Birmingham NHS Foundation Trust |
| Saralaya | D | Bradford Teaching Hospitals NHS Foundation Trust |
| Saratzis | A | University of Leicester |
| Sargant | J | University of Leicester |
| Sarginson | C | York & Scarborough NHS Foundation Trust |
| Sass | T | University Hospital Southampton NHS Foundation Trust |
| Sattar | N | University of Glasgow |
| Saunders | K | Oxford University Hospitals NHS Foundation Trust |
| Saunders | R | University of Leicester |
| Saunders | P | Sheffield Teaching NHS Foundation Trust |
| Saunders | L C | University of Sheffield |
| Savill | H | Tameside and Glossop Integrated Care NHS Foundation Trust |
| Saxon | W | Betsi Cadwallader University Health Board |
| Sayer | A | Newcastle upon Tyne Hospitals NHS Foundation Trust |
| Schronce | J | Imperial College Healthcare NHS Trust |
| Schwaeble | W | University of Cambridge |
| Scott | J T | MRC - University of Glasgow Centre for Virus Research |
| Scott | K | NHS Greater Glasgow and Clyde Health Board |
| Selby | N | Nottingham University |
| Semple | M G | Liverpool University Hospitals NHS Foundation Trust |
| Sereno | M | University Hospitals of Leicester NHS Trust |
| Sewell | T A | Sherwood Forest Hospitals NHS Foundation Trust |
| Shah | A M | King’s College Hospital NHS Foundation Trust |
| Shah | K | Diabetes UK |
| Shah | P | Royal Brompton and Harefield Clinical Group, Guy’s and St Thomas’ NHS Foundation trust. |
| Shankar-Hari | M | University of Edinburgh |
| Sharma | M | University of Leicester |
| Sharpe | C | King's College London |
| Sharpe | M | Oxford University Hospitals NHS Foundation Trust |
| Shashaa | S | East Cheshire NHS Trust |
| Shaw | A | Airedale NHS Foundation Trust |
| Shaw | K | Nottingham University Hospitals NHS Trust |
| Shaw | V | Liverpool University Hospitals NHS Foundation Trust |
| Sheikh | A | NHS Lothian & University of Edinburgh |
| Shelton | S | Sherwood Forest Hospitals NHS Foundation Trust |
| Shenton | L | Airedale NHS Foundation Trust |
| Shevket | K | King’s College Hospital NHS Foundation Trust |
| Shikotra | A | University Hospitals of Leicester NHS Trust |
| Short | J | University Hospital Birmingham NHS Foundation Trust |
| Siddique | S | St George’s University Hospitals NHS Foundation Trust |
| Siddiqui | S | University Hospitals of Leicester NHS Trust |
| Sidebottom | J | Sheffield Teaching NHS Foundation Trust |
| Sigfrid | L | University of Oxford |
| Simons | G | University of Southampton |
| Simpson | J | Newcastle upon Tyne Hospitals NHS Foundation Trust |
| Simpson | N | Imperial College Healthcare NHS Trust |
| Singapuri | A | University Hospitals of Leicester NHS Trust |
| Singh | C | Royal Free London NHS Foundation Trust |
| Singh | S | Royal Brompton and Harefield Clinical Group, Guy’s and St Thomas’ NHS Foundation trust. |
| Singh | S J | University Hospitals of Leicester NHS Trust |
| Sissons | D | Sherwood Forest Hospitals NHS Foundation Trust |
| Skeemer | J | University Hospitals of Leicester NHS Trust |
| Slack | K | Sherwood Forest Hospitals NHS Foundation Trust |
| Smith | A | NHS Lanarkshire |
| Smith | D | Imperial College London |
| Smith | S | Sherwood Forest Hospitals NHS Foundation Trust |
| Smith | J | Sheffield Teaching NHS Foundation Trust |
| Smith | L | Sheffield Teaching NHS Foundation Trust |
| Soares | M | University Hospitals of Leicester NHS Trust |
| Solano | T S | Guy’s and St Thomas’ NHS Foundation Trust |
| Solly | R | East Kent Hospitals University NHS Foundation Trust |
| Solstice | AR | NHS Tayside |
| Soulsby | T | University Hospital Birmingham NHS Foundation Trust |
| Southern | D | Betsi Cadwallader University Health Board |
| Sowter | D | Sherwood Forest Hospitals NHS Foundation Trust |
| Spears | M | University of Glasgow |
| Spencer | L G | University of Liverpool |
| Speranza | F | King’s College Hospital NHS Foundation Trust |
| Stadon | L | North Bristol NHS Trust |
| Stanel | S | University of Manchester |
| Steele | N | Sheffield Teaching NHS Foundation Trust |
| Steiner | M | University of Leicester |
| Stensel | D | Loughborough University |
| Stephens | G | Sheffield Teaching NHS Foundation Trust |
| Stephenson | L | Harrogate and District NHD Foundation Trust |
| Stern | M | Whittington Health NHS Trust |
| Stewart | I | Imperial College London |
| Stimpson | R | Sheffield Teaching NHS Foundation Trust |
| Stockdale | S | Manchester University NHS Foundation Trust |
| Stockley | J | University Hospital Birmingham NHS Foundation Trust |
| Stoker | W | Gateshead NHS Trust |
| Stone | R | Belfast Health & Social Care Trust & Queen's University Belfast |
| Storrar | W | Hampshire Hospitals NHS Foundation Trust |
| Storrie | A | Aneurin Bevan University Health Board |
| Storton | K | Bradford Teaching Hospitals NHS Foundation Trust |
| Stringer | E | University Hospitals of Leicester NHS Trust |
| Strong-Sheldrake | S | Salisbury NHS Foundation Trust |
| Stroud | N | Cwm Taf Morgannwg University Health Board |
| Subbe | C | Betsi Cadwallader University Health Board |
| Sudlow | C L | University of Edinburgh |
| Suleiman | Z | University Hospital Birmingham NHS Foundation Trust |
| Summers | C | University of Cambridge |
| Summersgill | C | Salford Royal NHS Foundation Trust |
| Sutherland | D | NHS Tayside |
| Sykes | D L | Hull University Teaching Hospitals NHS Trust |
| Sykes | R | NHS Greater Glasgow and Clyde Health Board |
| Talbot | N | Oxford University Hospitals NHS Foundation Trust |
| Tan | A L | Leeds Teaching Hospitals |
| Tarusan | L | Imperial College Healthcare NHS Trust |
| Tavoukjian | V | St George’s University Hospitals NHS Foundation Trust |
| Taylor | A | Hywel Dda University Health Board |
| Taylor | C | University of Leicester |
| Taylor | J | Cambridge University Hospitals NHS Foundation Trust |
| Te | A | King’s College Hospital NHS Foundation Trust |
| Tedd | H | Newcastle upon Tyne Hospitals NHS Foundation Trust |
| Tee | CJ | NHS Tayside |
| Teixeira | J | St George’s University Hospitals NHS Foundation Trust |
| Tench | H | Hywel Dda University Health Board |
| Terry | S | University of Leicester |
| Thackray-Nocera | S | Hull University Teaching Hospitals NHS Trust |
| Thaivalappil | F | Swansea Bay University Health Board |
| Thamu | B | Sheffield Teaching NHS Foundation Trust |
| Thickett | D | University of Birmingham |
| Thomas | C | Swansea Bay University Health Board |
| Thomas | D C | Imperial College Healthcare NHS Trust |
| Thomas | S | Newcastle upon Tyne Hospitals NHS Foundation Trust |
| Thomas | A K | Nottingham University Hospitals NHS Trust |
| Thomas-Woods | T | Cwm Taf Morgannwg University Health Board |
| Thompson | T | University Hospital Birmingham NHS Foundation Trust |
| Thompson | A A R | Sheffield Teaching NHS Foundation Trust |
| Thornton | T | University Hospitals of Leicester NHS Trust |
| Thorpe | M | University of Edinburgh |
| Thwaites | R S | Imperial College London |
| Tilley | J | Somerset NHS Foundation Trust |
| Tinker | N | Sheffield Teaching NHS Foundation Trust |
| Tiongson | G F | London North West University Healthcare NHS Trust |
| Tobin | M | University Hospitals of Leicester NHS Trust |
| Tomlinson | J | Shropshire Community Health NHS Trust |
| Tong | C | University of Leicester |
| Toshner | M | Cambridge University Hospitals NHS Foundation Trust |
| Touyz | R | Institute of Cardiovascular & Medical Sciences, University of Glasgow |
| Tripp | K A | Liverpool University Hospitals NHS Foundation Trust |
| Tunnicliffe | E | Oxford University Hospitals NHS Foundation Trust |
| Turnbull | A | York & Scarborough NHS Foundation Trust |
| Turner | E | University of Leicester |
| Turner | S | Sherwood Forest Hospitals NHS Foundation Trust |
| Turner | V | Tameside and Glossop Integrated Care NHS Foundation Trust |
| Turner | K | Sheffield Teaching NHS Foundation Trust |
| Turney | S | East Kent Hospitals University NHS Foundation Trust |
| Turtle | L | Liverpool University Hospitals NHS Foundation Trust |
| Turton | H | Sheffield Teaching NHS Foundation Trust |
| Ugoji | J | The Great Western Hospital Foundation Trust |
| Ugwuoke | R | Salford Royal NHS Foundation Trust |
| Upthegrove | R | University of Birmingham |
| Valabhji | J | Imperial College London |
| Ventura | M | University Hospital Birmingham NHS Foundation Trust |
| Vere | J | Tameside and Glossop Integrated Care NHS Foundation Trust |
| Vickers | C | Somerset NHS Foundation Trust |
| Vinson | B | University of Liverpool |
| Wade | E | Leeds Teaching Hospitals |
| Wade | P | Sheffield Teaching NHS Foundation Trust |
| Wain | L V | University Hospitals of Leicester NHS Trust |
| Wainwright | T | Somerset NHS Foundation Trust |
| Wajero | L O | Liverpool University Hospitals NHS Foundation Trust |
| Walder | S | University Hospital Birmingham NHS Foundation Trust |
| Walker | S | Sheffield Teaching NHS Foundation Trust |
| Wall | E | University College London Hospital |
| Wallis | T | University Hospital Southampton NHS Foundation Trust |
| Walmsley | S | University of Edinburgh |
| Walsh | J A | Royal Brompton and Harefield Clinical Group, Guy’s and St Thomas’ NHS Foundation trust. |
| Walsh | S | Imperial College London |
| Warburton | L | Shropshire Community Health NHS Trust |
| Ward | T J C | University Hospitals of Leicester NHS Trust |
| Warwick | K | Kettering General Hospital NHS Trust |
| Wassall | H | East Cheshire NHS Trust |
| Waterson | S | North Bristol NHS Trust |
| Watson | E | London North West University Healthcare NHS Trust |
| Watson | L | Cambridge University Hospitals NHS Foundation Trust |
| Watson | J | Sheffield Teaching NHS Foundation Trust |
| Welch | C | University Hospital Birmingham NHS Foundation Trust |
| Welch | H | North Bristol NHS Trust |
| Welsh | B | NHS Lanarkshire |
| Wessely | S | King's College London |
| West | S | Newcastle upon Tyne Hospitals NHS Foundation Trust |
| Weston | H | East Kent Hospitals University NHS Foundation Trust |
| Wheeler | H | University Hospital Southampton NHS Foundation Trust |
| White | S | Kettering General Hospital NHS Trust |
| Whitehead | V | Betsi Cadwallader University Health Board |
| Whitney | J | King's College London |
| Whittaker | S | Salford Royal NHS Foundation Trust |
| Whittam | B | Leeds Teaching Hospitals |
| Whitworth | V | Sherwood Forest Hospitals NHS Foundation Trust |
| Wight | A | Wirral University Teaching Hospital |
| Wild | J | University of Sheffield |
| Wilkins | M | Imperial College London |
| Wilkinson | D | University of Birmingham |
| Williams | B | University College London |
| Williams | N | Hampshire Hospitals NHS Foundation Trust |
| Williams | J | Cardiff and Vale University Healthy Board |
| Williams-Howard | S A | Liverpool University Hospitals NHS Foundation Trust |
| Willicombe | M | Imperial College London |
| Willis | G | Aneurin Bevan University Health Board |
| Willoughby | J | University College London |
| Wilson | A | Gateshead NHS Trust |
| Wilson | D | University Hospital Birmingham NHS Foundation Trust |
| Wilson | I | Sheffield Teaching NHS Foundation Trust |
| Window | N | Leeds Teaching Hospitals |
| Witham | M | Newcastle upon Tyne Hospitals NHS Foundation Trust |
| Wolf-Roberts | R | Hywel Dda University Health Board |
| Wood | C | Imperial College Healthcare NHS Trust |
| Woodhead | F | University Hospitals of Leicester NHS Trust |
| Woods | J | Leeds Teaching Hospitals |
| Wootton | D G | Liverpool University Hospitals NHS Foundation Trust |
| Wormleighton | J | University of Leicester |
| Worsley | J | Cambridge University Hospitals NHS Foundation Trust |
| Wraith | D | University of Birmingham |
| Wrey Brown | C | Hampshire Hospitals NHS Foundation Trust |
| Wright | C | Hull University Teaching Hospitals NHS Trust |
| Wright | L | University of Nottingham |
| Wright | S | Newcastle upon Tyne Hospitals NHS Foundation Trust |
| Wyles | J | Liverpool University Hospitals NHS Foundation Trust |
| Wynter | I | Sherwood Forest Hospitals NHS Foundation Trust |
| Xie | C | University of Oxford, Division of Cardiovascular Medicine |
| Xu | M | University College London |
| Yasmin | N | Imperial College Healthcare NHS Trust |
| Yasmin | S | University Hospital Birmingham NHS Foundation Trust |
| Yates | T | University Hospitals of Leicester NHS Trust |
| Yip | K P | University Hospital Birmingham NHS Foundation Trust |
| Young | B | DUK \| NHS Digital, Salford Royal Foundation Trust |
| Young | S | Aneurin Bevan University Health Board |
| Young | A | Newcastle upon Tyne Hospitals NHS Foundation Trust |
| Yousuf | A J | University Hospitals of Leicester NHS Trust |
| Zawia | A | Sheffield Teaching NHS Foundation Trust |
| Zeidan | L | The Rotherham NHS Foundation Trust |
| Zhao | B | University of Leicester |
| Zongo | O | Barts Health NHS Trust |
| Zheng | B | University of Edinburgh |
